# Cross-Institutional European Evaluation and Validation of Automated Multilabel Segmentation for Acute Intracerebral Hemorrhage and Complications

**DOI:** 10.1101/2024.08.27.24312653

**Authors:** Jawed Nawabi, Georg Lukas Baumgärtner, Sophia Schulze-Weddige, Andrea Dell’Orco, Andrea Morotti, Federico Mazzacane, Helge Kniep, Frieder Schlunk, Maik FH Böhmer, Burak Han Akkurt, Tobias Orth, Weissflog Jana Sofie, Maik Schumann, Peter B. Sporns, Michael Scheel, Uta Hanning, Jens Fiehler, Tobias Penzkofer

## Abstract

**Purpose:** To evaluate a nnU-Net-based deep learning for automated segmentation of intracerebral hemorrhage (ICH), intraventricular hemorrhage (IVH), and perihematomal edema (PHE) on noncontrast CT scans.

**Materials and Methods:** Retrospective data from acute ICH patients admitted at four European stroke centers (2017-2019), along healthy controls (2022-2023), were analyzed. nnU-Net was trained (n=775) using a 5-fold cross-valiadtion approach, tested (n=189), and seperatly validated on internal (n=121), external (n=169), and diverse ICH etiologies (n=175) datasets. Interrater-validated ground truth served as the reference standard. Lesion detection, segmentation, and volumetric accuracy were measured, alongside time efficiency versus manual segmentation.

**Results:** Test set results revealed high nnU-Net accuracy (median Dice Similartiy Coefficient (DSC): ICH 0.91, IVH 0.76, PHE 0.71) and volumetric correlation (ICH, IVH: r=0.99; PHE: r=0.92). Sensitivities were high (ICH, PHE: 99%; IVH: 97%), with IVH detection specificities and sensitivities >90% for volumes up to 0.2 ml. Anatomical-specific metrics showed higher performance for lobar and deep hemorrhages (median DSC 0.90 and 0.92, respectively) and lower for brainstem (median DSC 0.70). Concurrent hemorrhages did not affect accuracy, p> 0.05. Across validation sets, segmentation precision was consistent, especially for ICH (median DSC 0.85-0.90), with PHE slightly lower (median DSC 0.61-0.66) and IVH best in the second and third set (median DSC 0.80). Average processing time was 18.2 seconds versus 18.01 minutes manually.

**Conclusion:** The nnU-Net provides reliable, time-efficient ICH, IVH, and PHE segmentation, validated across various clinical settings, with excellent anatomical-specific performance for lobar and deep hemorrhages. It shows promise for enhancing clinical workflow and research initiatives.

## 1 Introduction

Acute intracerebral hemorrhage (ICH), a major cause of stroke mortality, requires neuroimaging for etiology investigation and management, with the volume quantification of ICH and secondary complications like intraventricular hemorrhage (IVH) and perihematomal edema (PHE) being crucial for clinical decisions.^1,2,3,4,5^ The complexity of ICH demands sophisticated analysis, now enhanced by Deep Learning segmentation techniques.^6,7^ However, efforts to comprehensively capture thisphenotype of ICH are rare, as prior research has mainly focused only on ICH segmentation, PHE segmentation, or a combination of the two.^8,9,10^ We developed a neural network trained on CT images from three European centers, validated across datasets including diverse ICH etiologies, to precisely quantify and detect ICH, IVH, and PHE, as detailed in our methodology.

## 2 Methods and Material

### 2.1 Study Design

This retrospective multicenter study received approval from the ethics committees of Charité Berlin, Germany (protocol number EA1/035/20), University Medical-Center Hamburg, Germany (protocol number WF-054/19), University Hospital Munster, Germany [2017-233-f-S], and and IRCCS Mondino Foundation Pavia, Italy (protocol number 20190099462). The institutional review boards waived the requirement for written informed consent. All protocols and procedures adhered to the Declaration of Helsinki. Given the study’s retrospective nature, patient consent was not required.

### 2.2 Data Collection and Patient Demographics

Our retrospective analysis encompassed patients with acute ICH from four major stroke centers in Germany and Italy. A control group was established, comprising baseline non-contrast NCCT scans from healthy individuals over the age of 18, recruited from Charité Berlin, Germany, between 2022-2023. A comprehensive description of the inclusion and exclusion criteria is provided in the Supplementary Material.

### 2.3 Image Acquisitions

The NCCT baseline scans were acquired in adherence to the individual protocols of the contributing centers, utilizing a variety of CT scanners. Specifically, data was collected using 14 distinct scanner models across 4 manufacturers, with detailed information provided in Supplementary Table 1.

### 2.4 Imaging Analysis and Image Quality Assessment

Quantitative and qualitative analyses of CT scans were performed to assess IVH and hemorrhage location, using detailed imaging annotation and manual region segmentation. A thorough explanation of the image analysis, quality assessment protocols, and the exclusion of images based on defined criteria in provided in the Supplementary Material.

### 2.5 Deep Learning Network Architecture

We utilized the adaptable nnU-Net framework for semantic segmentation of biomedical images, characterized by its dynamic adjustment to datasets and a detailed U-Net-like architecture, with specifics on its encoder-decoder structure and parameter groups depicted in **Figure 1** and further elaborated in the Supplementary Material.^13,14^

**Figure 1:**
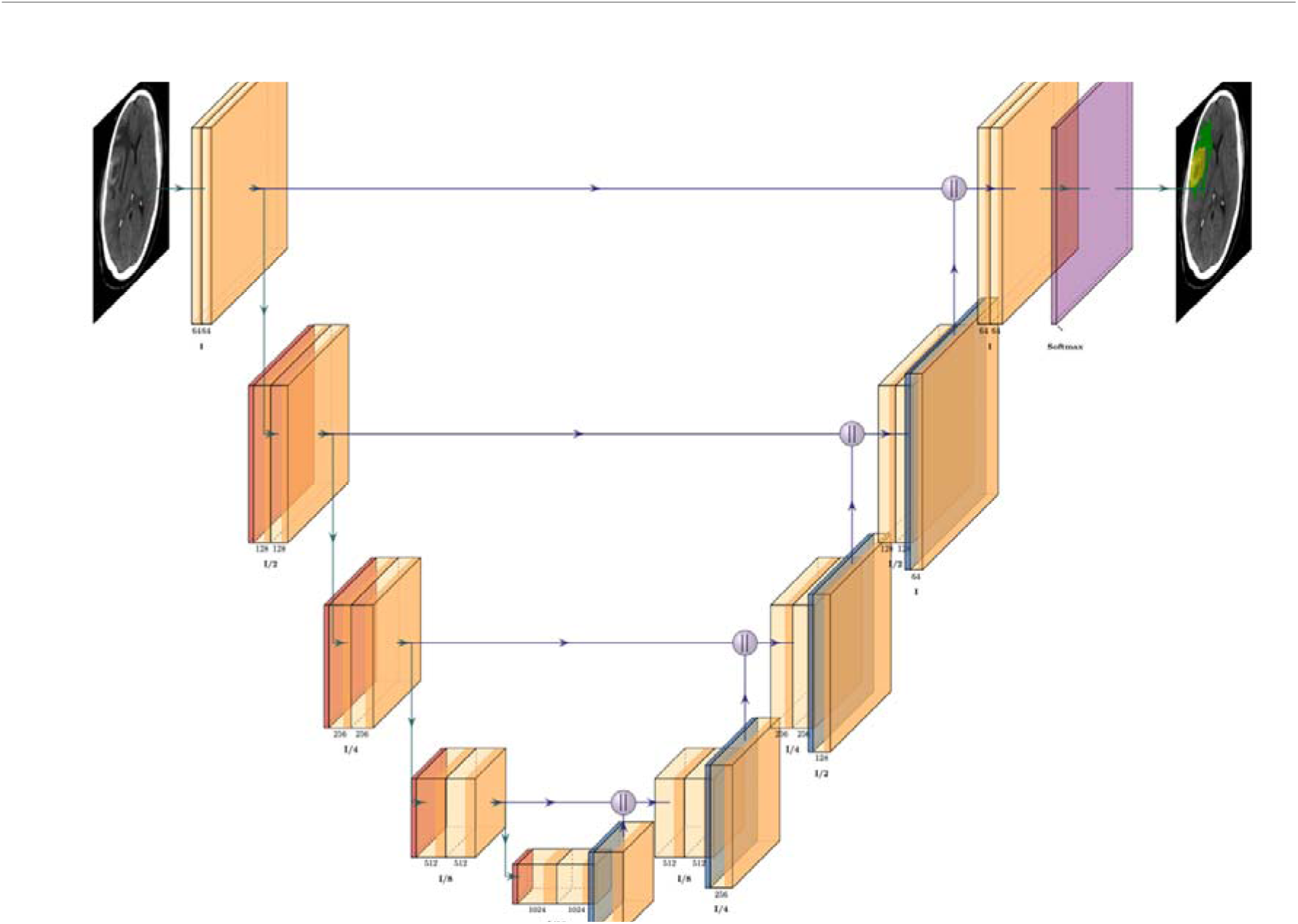
Deep Learning Architecture of the customized 3D U-Net in the nnU-Net. *Legend:* Illustrated nnU-Net framework for automated segmentation of intracerebral hemorrhage (ICH), perihematomal edema (PHE), and intraventricular hemorrhage (IVH) involving several steps: **(1) Data Preprocessing:** The initial stage involves preprocessing of input data, which includes standardizing intensity and resolution to prepare for segmentation. **(2) Segmentation Mechanism:** Utilizing nnU-Net architecture, the algorithm employs a block that contains a 3x3x3 convolution, instance normalization, and ReLU activations for segmenting ICH, PHE, and IVH. **(3) Volumetric Quantification:** Following segmentation, the volumes of ICH, IVH, and PHE are calculated. **(4) Network Configuration and Training:** nnU-Net fully automatically configures and trains the entire segmentation pipeline, tailored to the dataset’s characteristics. (5) Adaptive Scaling: The network can adjust to the input patch size, allowing up to seven down-sampling and up-sampling operations, potentially increasing the feature representation significantly at the bottleneck layer. **(6) Softmax Function:** Finally, a softmax function is applied to the output layers to produce probabilistic maps, which assist in distinguishing between the different types of hemorrhagic lesions.

### 2.6 Experimental Protocol

**2.6.1 Training and Testing**

Training involved subjects from Charité Berlin, University Medical-Center Hamburg, and IRCCS Mondino Foundation Pavia with 20% of data reserved for testing and 80% undergoing 5-fold cross-validation to boost model robustness. Each fold contributed to training and validation, with final predictions combining all folds. The test set remained separate, and performance was assessed by comparing predicted and ground truth masks, as well as segmentation times; details are in the Supplementary Material.

#### 2.6.2 Validation

Our segmentation network’s robustness was tested using three datasets: the Internal Validation Set (Validation Set #1) from University Medical Center Hamburg-Eppendorf, Germany, the External Validation Set (Validation Set #2) from University Hospital Munster, Germany for generalizability, and the Diverse Validation Set from Charité Berlin (Validation Set #3) covering various ICH subtypes for broad-spectrum assessment. Additional details are available in the Supplementary Material.

#### 2.6.3 Efficiency of Automated Segmentation

A comparative analysis of the time efficiency between manual segmentation performed by a trained reader and our automated deep learning segmentation network was conducted. This analysis involved calculating the aggregate time required for delineating ICH, PHE, and IVH on a per-case basis utilizing both methodologies.

### 2.7 Training and Loss functions

Models underwent training for 1000 epochs, with an initial learning rate of 0.01 and batches of 12 images. We utilized a combined loss function of Dice loss and categorical cross-entropy, the latter being a standard for multi-class problems, to compute the divergence of output probabilities from true values (details in Supplementary Material).^15^ Loss optimization leveraged stochastic gradient descent with the same learning rate specified.^16^ The entire experiment was conducted using Python (v3.8.10)^17^ and nnU-Net (v2.0)^18^ using various NVIDIA GPUs, including Quadro RTX 8000, A100 and H100.^19^ The code used for training and inference of the deep learning systems is available online from the original github repository.^18^

### 2.7 Performance Evaluation Metric

Model and volume estimation performance were measured using Dice Similarity Coefficient (DSC), Absolute Volume Error (AVE), absolute relative volume difference (ARVD), and correlation coefficients. ^20,21,22^ Automated detection accuracy was benchmarked using traditional metrics. Subgroup analyses focused on lesions >1ml for clinical relevance and cases with secondary bleedings, evaluating accuracy with DSC and Pearson correlation.^23,24^ Further methodological details, including formulas and subgroup analyses, are provided in the Supplementary Material.

### 2.8 Analysis Time

Time efficiency was assessed by measuring the duration of manual segmentation per case and comparing it to the network’s automated segmentation from initiation to the delivery of results. Statistical analyses were performed using Scikit-learn library (Python v3.8.10; Python Software Foundation).^17,25^

### 2.9 A Web-based User-Interface for Radiological Report

We developed a web-based interface, the ICH-Viewer, for segment visualization and volume reporting, utilizing Cornerstone.js for DICOM image handling and Flask for UI construction; **Figure 2** displays the ICH-Viewer Interface, with further technical details available in the Supplementary Material.^26,27^

**Figure 2:**
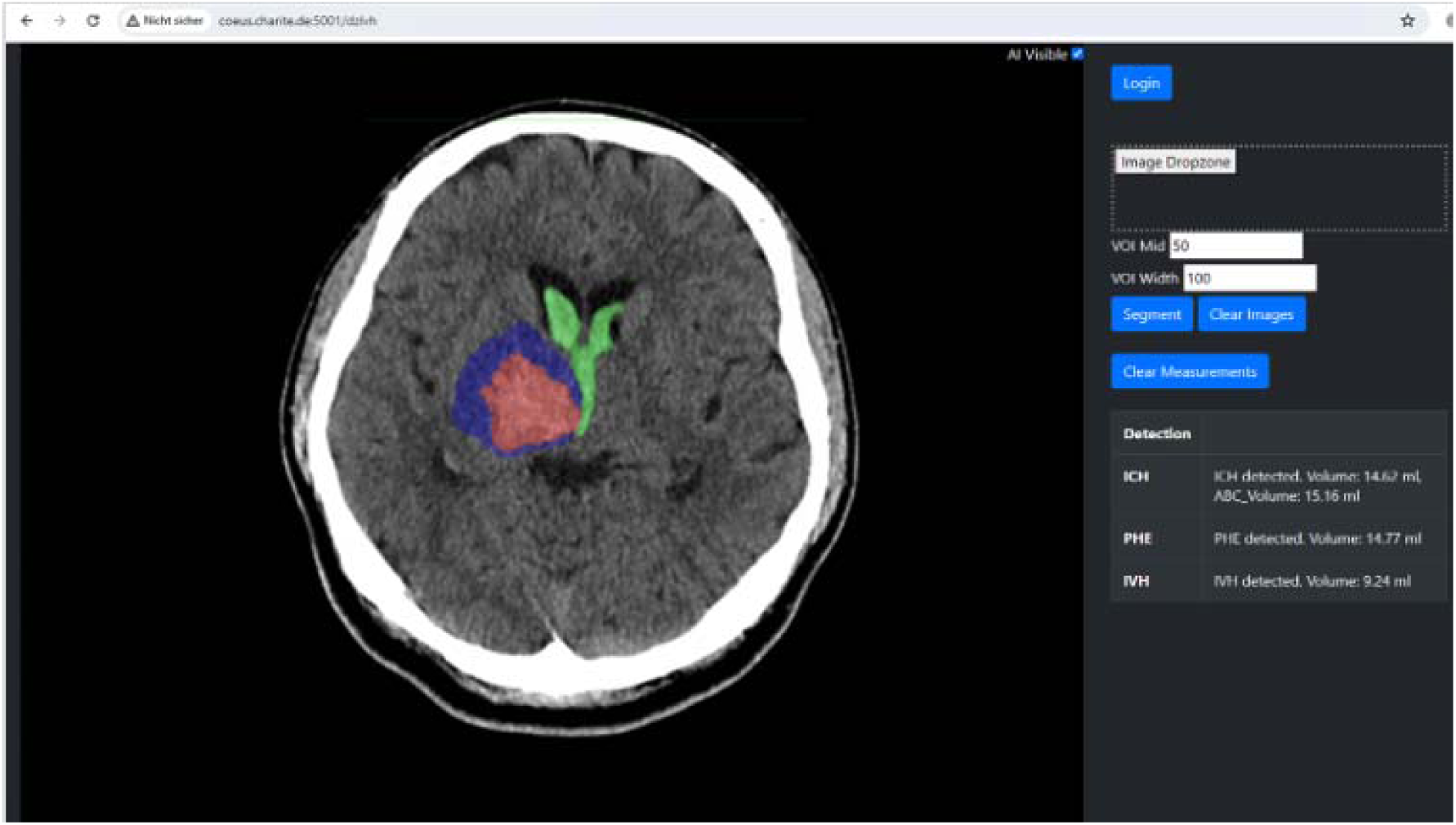
ICH-Viewer Interface: Integrative Display for Image Analysis and Volume Reporting. *Legend:* Interface Snapshot: Automated segmentation visualization for intracerebral hemorrhage (red) in the basal ganglia with adjacent perihematomal edema (blue) and intraventricular involvement (green). Side panels feature a DICOM or Nifti dropzone for ordering or clearing the segmentation and a dedicated viewport displaying automated volumetric reports alongside volumes generated via the ABC/2 Method for ICH assessment. ICH, intracerebral hemorrhage; IVH, intraventricular hemorrhage; PHE, perihematomal hemorrhage.

## Results

### Characteristics of the study cohort

The characteristics of the training and test cohorts were generally well-matched, ensuring consistency for model validation. However, there were differences in gender distribution, GCS score at baseline, and IVH volume, which were more pronounced in the test cohort as detailed in **Table 1**. Validation Set #1 included 121 patients, Set #2 comprised 169 patients, and Set #3 contained 175 patients; detailed patient and imaging characteristics are provided in **Table 1**.

**Table 1:**
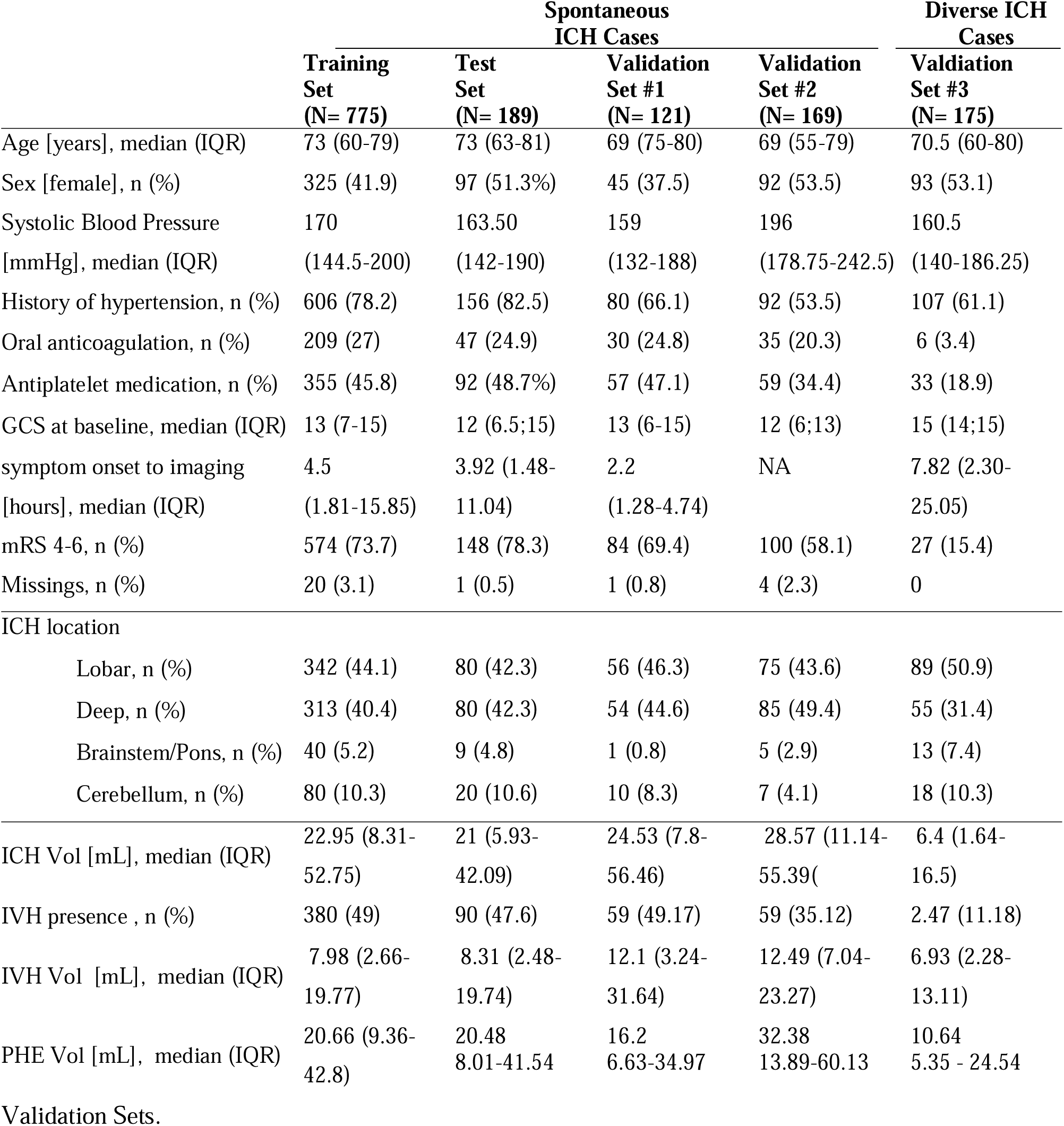
Patient and Imaging Characteristics Across Training, Test, and Independent. *Legend:* GCS, Galsgow Coma Scale; ICH, intracerebral hemorrhage; IQR, interquartile range; IVH, intraventricular hemorrhage; mRS, modified Rankin Scale; PHE, perihematomal edema; SD, standard deviation; Vol, volume.

### Performance of lesion segmentation, detection and volume quantification

**Table 2** presents the comparative performance of lesion segmentation, detection and volume quantification on the test set against the reference standard, which was established through repeated ratings and manual measurements. For ICH, the network achieved a median DSC of 0.91 (±0.01) against the reference standard, p-value of 0.1962. The volume difference in ICH detection was minimal, at 5% (±10%) with an absolute volume difference of -0.62 (±0.65) ml, showing close alignment the reference standard. We investigated where this underestimation occurred in a Bland-Altman plot in **Figure 3, A**, which displayed some variability increases in cases with an ICH volume > 100mL. Nonetheless, the majority of the automatic volumes fell within 10 mL of the manual volumes. In line with this, the network demonstrated high precision in volume estimation, with a Pearson correlation of 0.99, and a robust correlation with DSC, as depicted in the scatterplot in **Figure 4, A**. The illustrative cases featured in **Figure 5-6**, further detail the network’s robust performance in handling complex hemorrhages. Detection accuracy and sensitivity reached a consistent high of 99% (±1%), with performance demonstrated by cases identifying even small ICHs, as shown in **Supplementary Figure 1, A**.

**Figure 3:**
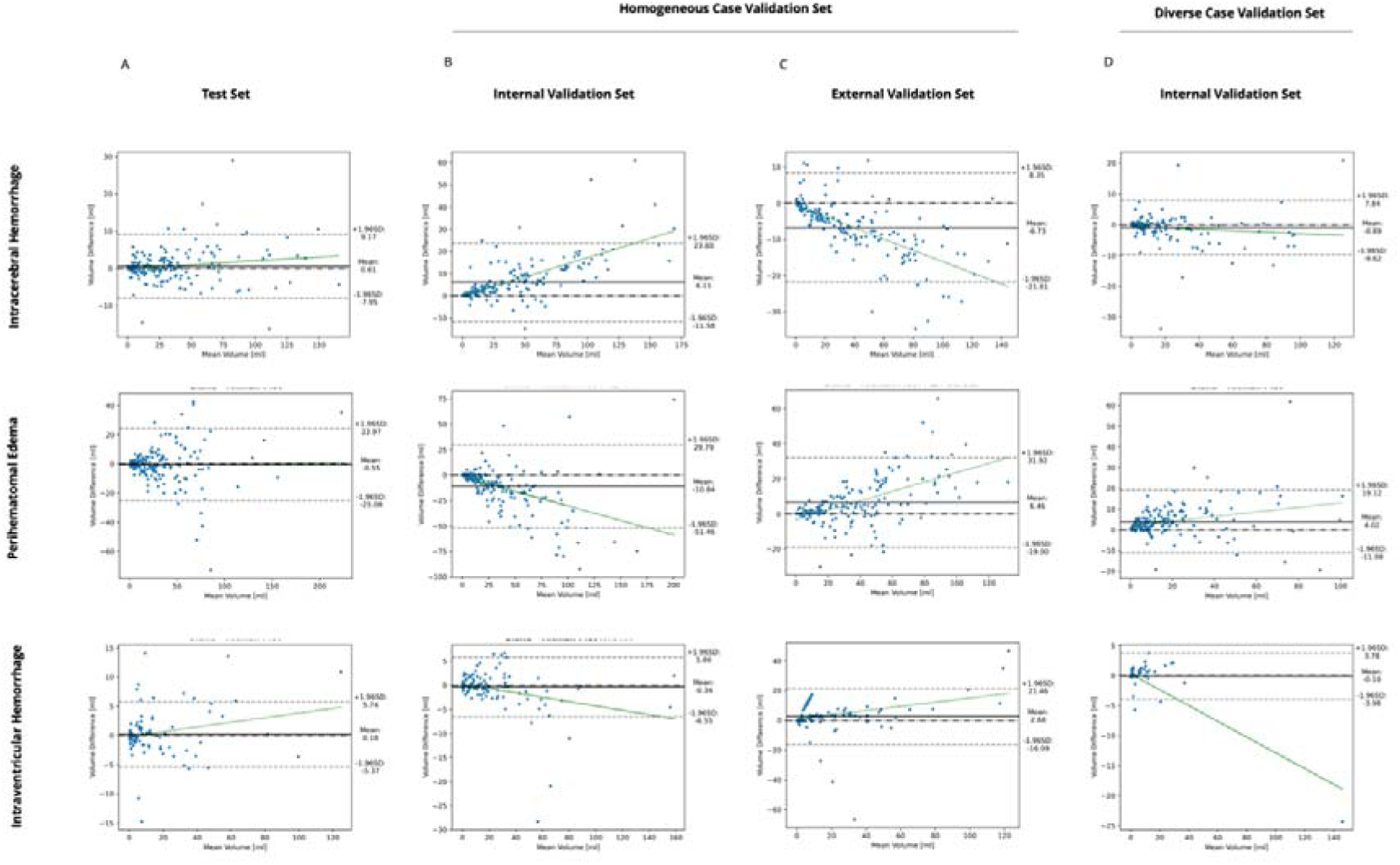
Bland-Altman Analysis for Model Performance on Test Set Versus Homogenous and Diverse Internal and External Validation Sets. *Legend:* Bland-Altman Analysis for assessing the concordance between automated segmentation volumes and the established ground truth for intracerebral hemorrhage (ICH; upper row), perihematomal edema (PHE; mid row), and intraventricular hemorrhage (IVH; bottom row). The x-axis represents the mean volume derived from automated segmentation, while the y-axis depicts the volume difference. Where, positive values indicate underestimation by the automated method relative to the manual gold standard, whereas negative values suggest overestimation. The central solid line denotes the mean absolute volume deviation from the true volume, with the surrounding dotted lines representing the bounds of one standard deviation. Panel A illustrates the analysis within the test set, Panel B and C demonstrate the findings in homogeneous validation sets comprising solely spontaneous ICH cases (internal and external sets, respectively), and Panel D showcases the analysis within a heterogeneous validation set, encompassing a broad range of ICH etiologies.

**Figure 4:**
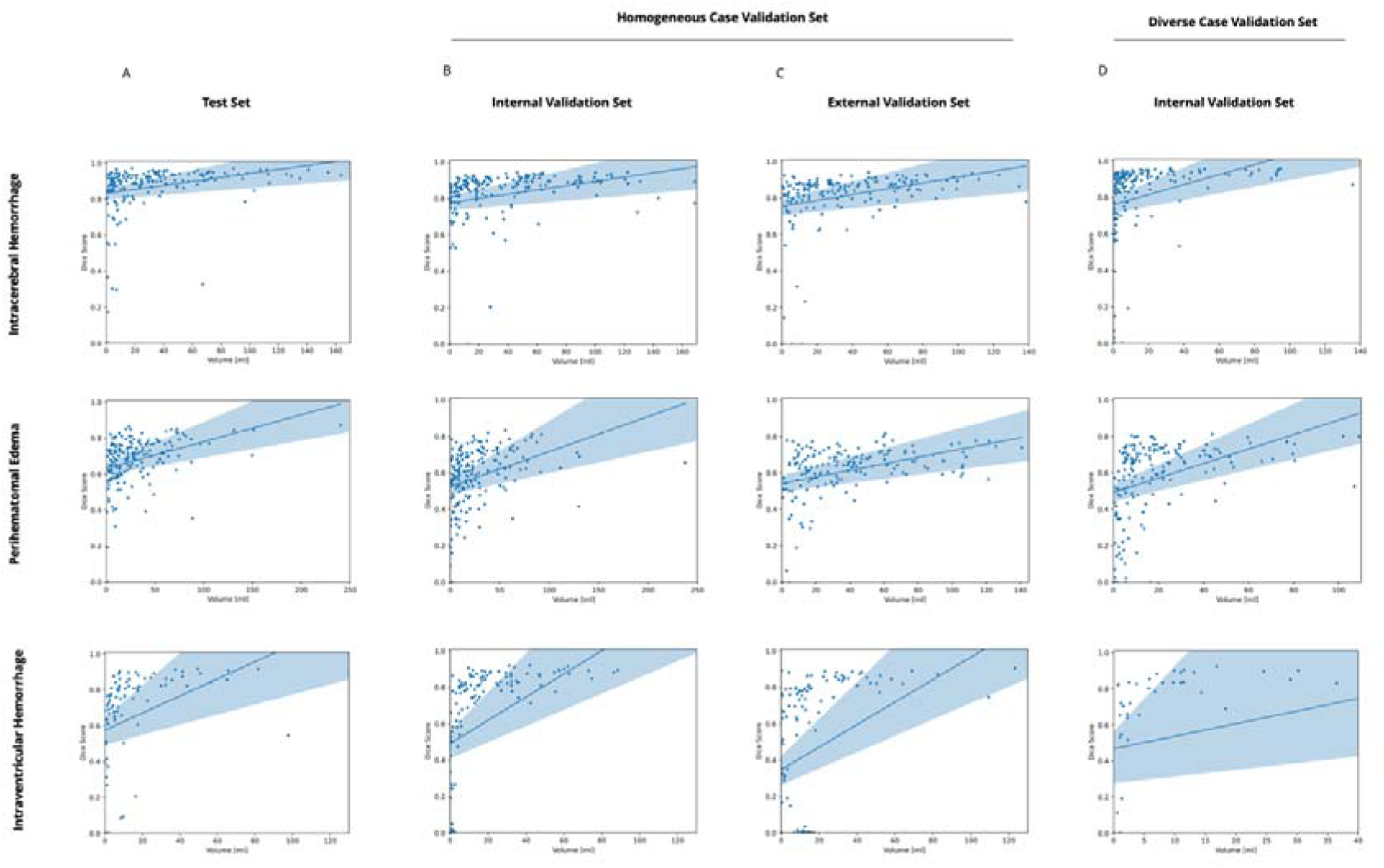
Correlation Scatter Plot Analysis for Model Performance on Test Set Versus Homogenous and Diverse Internal and External Validation Sets. *Legend:* Scatter plots illustrating the correlation between automated segmentation volumes and ground truth across various data sets. Each point represents the automated volume versus the manual (ground truth) volume for intracerebral hemorrhage (ICH), perihematomal edema (PHE), and intraventricular hemorrhage (IVH). Panel A focuses on the test set, while Panels B and C depict homogeneous validation sets for spontaneous ICH cases—internal and external, respectively. Panel D examines a heterogeneous validation set covering a range of ICH causes. The trend lines indicate the direction and strength of the correlation, providing insight into the predictive accuracy of the segmentation model. The shaded blue region around the trend line in the scatter plot represents the 95% confidence interval.

**Figure 5:**
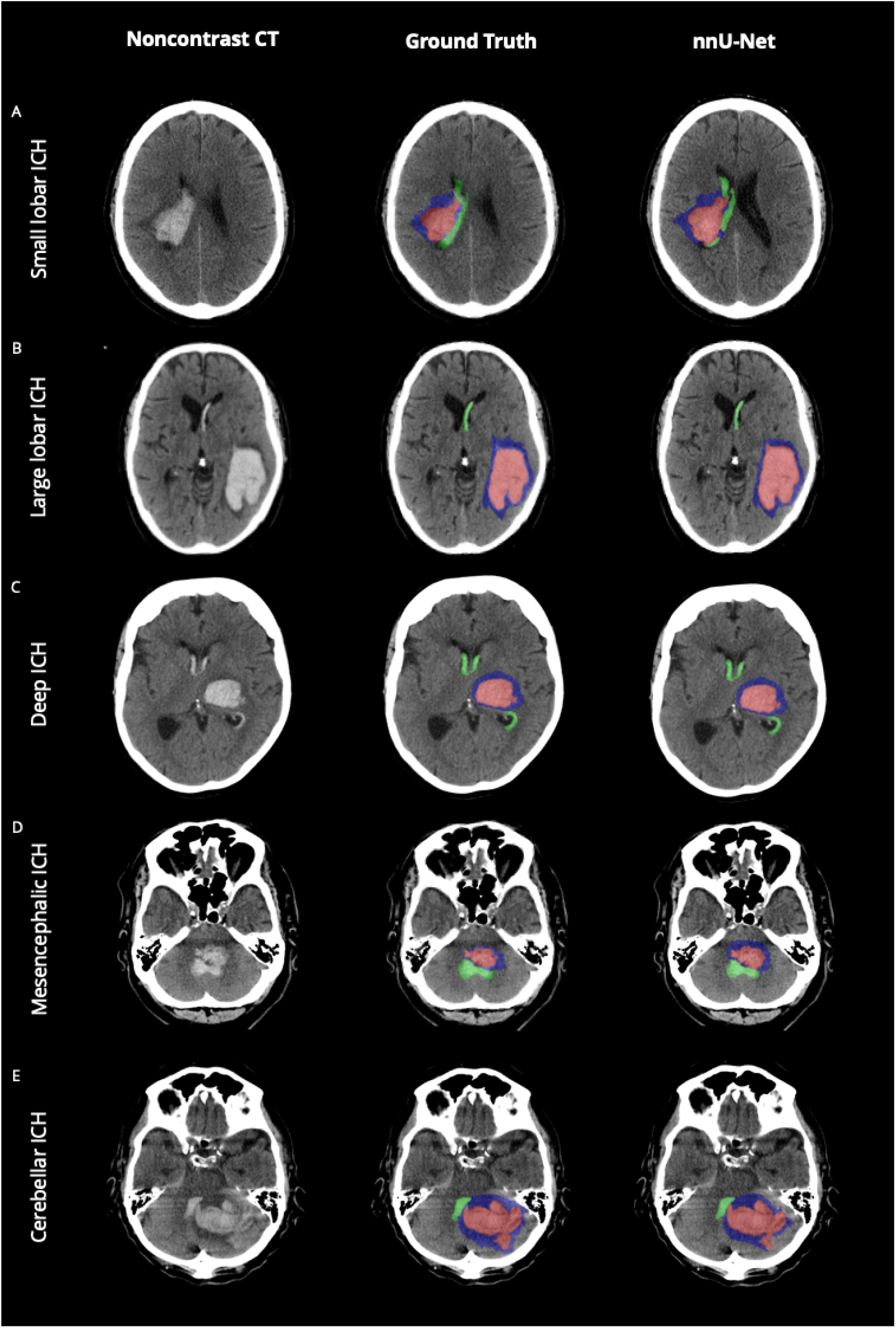
Comparative Visualization of Model Predictions versus Ground Truth across various Anatomical Contexts. *Legend:* Noncontrast Computed Tomography scans with the original data (left column), ground truth segmentations (mid column) and the network’s predicted segmentations (nnU-Net, right column) for different anatomical contexts. Each row represents a distinct anatomical context, illustrating the model’s ability to identify and segment key pathological features. The color-coding is as follows: intracerebral hemorrhage (ICH) is highlighted in red; intraventricular hemorrhage extension (IVH) is marked in green; and perihematomal edema (PHE) is depicted in blue.

**Figure 6:**
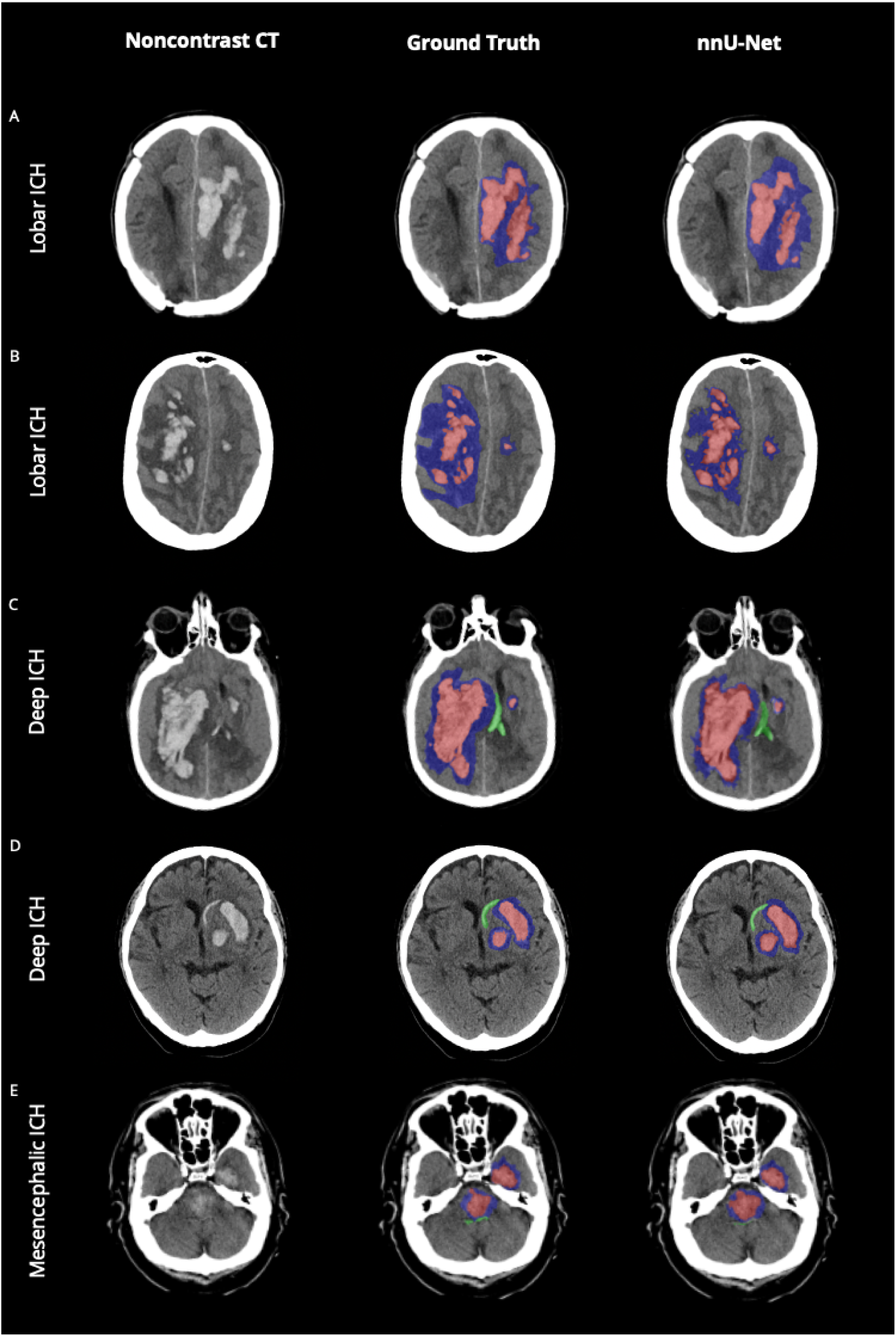
Comparative Visualization of Model Predictions versus Ground Truth across Complex Hematoma Configurations. *Legend:* Noncontrast Computed Tomography scans with the original data (left column), ground truth segmentations (mid column) and the network’s predicted segmentations (nnU-Net, right column) for different anatomical contexts. Each row represents a distinct anatomical context, illustrating the model’s ability to identify and segment key pathological features. The color-coding is as follows: intracerebral hemorrhage (ICH) is highlighted in red; intraventricular hemorrhage extension (IVH) is marked in green; and perihematomal edema (PHE) is depicted in blue.

**Table 2:** Evaluation of nnU-Net Framework: Lesion Segmentation, Volume Estimation, and Detection Accuracy Across the Test Data Set in Comparison with the Ground Truth. *Legend:* Performance metrics (1) for segmentation evaluation, including Dice score analysis, represented through mean and median values, along with pairwise comparison among various datasets; (2) volume estimation accuracy, utilizing both relative and absolute volume discrepancies, complemented by a Pearson correlation assessment between actual ground truth (GT) volumes and those predicted by the AI; and (3) the efficacy of detection across all data subjects and for those above 1 mL. Additional segmentation metrics for IVH exceeding 1 mL are specifically delineated in a separate subsection. Intracerebral hemorrhage (ICH), perihematomal edema (PHE) and intraventricular hemorrhage (IVH).

| Lesion Type | Performance Metrics | Test Dataset | Reference |
| --- | --- | --- | --- |
| ICH Metrics | Mean DICE Score | $0.87 \pm 0.02$ | $0.83 \pm 0.05$ |
| | Median DICE Score | $0.91 \pm 0.01$ | $0.86 \pm 0.03$ |
|  | T-Test vs Test Dataset; p-value |  | 0.1962 |
| | Volume Difference in % | $5\% \pm 10\%$ | $7\% \pm 19\%$ |
| | Absolute Difference in ml | $-0.63 \pm 0.65$ | $0.76 \pm 1.64$ |
|  | Pearson Correlation Volume GT vs AI | 0.99 | 0.98 |
| | Detection Accuracy | $99\% \pm 1\%$ | $100\% \pm 0\%$ |
| | Detection Sensitivity | $99\% \pm 1\%$ | $100\% \pm 0\%$ |
|  | Detection Specificity | NA | NA |
| | Detection Accuracy Threshold of 1 ml | $97\% \pm 3\%$ | $100\% \pm 0\%$ |
| | Detection Sensitivity Threshold of 1 ml | $97\% \pm 3\%$ | $100\% \pm 0\%$ |
|  | Detection Specificity Threshold of 1 ml | NA | NA |
| PHE Metrics | Mean DICE Score | $0.67 \pm 0.02$ | $0.67 \pm 0.07$ |
| | Median DICE Score | $0.71 \pm 0.02$ | $0.71 \pm 0.04$ |
|  | T-Test vs Test Dataset; p-value |  | 0.7521 |
| | Volume Difference in % | $12\% \pm 9\%$ | $-10\% \pm 13\%$ |
| | Absolute Difference in ml | $0.57 \pm 1.84$ | $-4.32 \pm 5.42$ |
|  | Pearson Correlation Volume GT vs AI | 0.92 | 0.91 |
| | Detection Accuracy | $98\% \pm 2\%$ | $100\% \pm 0\%$ |
| | Detection Sensitivity | $99\% \pm 1\%$ | $100\% \pm 0\%$ |
|  | Detection Specificity | NA | NA |
| | Detection Accuracy Threshold of 1 ml | $97\% \pm 2\%$ | $100\% \pm 0\%$ |
| | Detection Sensitivity Threshold of 1 ml | $98\% \pm 2\%$ | $100\% \pm 0\%$ |
|  | Detection Specificity Threshold of 1 ml | NA | NA |
| IVH Metrics | Mean DICE Score | $0.51 \pm 0.07$ | $0.82 \pm 0.10$ |
| | Median DICE Score | $0.68 \pm 0.09$ | $0.84 \pm 0.04$ |
|  | T-Test vs Test Dataset; p-value |  | 0.001014 |
| | Volume Difference in % | $-1\% \pm 10\%$ | $11\% \pm 4\%$ |
| | Absolute Difference in ml | $-0.87 \pm 0.83$ | $-2.10 \pm 1.8$ |
|  | Pearson Correlation Volume GT vs AI | 0.99 | 1 |
| | Detection Accuracy | $82\% \pm 6\%$ | $100\% \pm 0\%$ |
| | Detection Sensitivity | $97\% \pm 4\%$ | $100\% \pm 0\%$ |
| | Detection Specificity | $70\% \pm 9\%$ | $100\% \pm 0\%$ |
| | Detection Accuracy Threshold of 1 ml | $90\% \pm 5\%$ | $100\% \pm 0\%$ |
| | Detection Sensitivity Threshold of 1 ml | $86\% \pm 8\%$ | $100\% \pm 0\%$ |
| | Detection Specificity Threshold of 1 ml | $93\% \pm 5\%$ | $100\% \pm 0\%$ |
| IVH Threshold Metrics (> 1ml) | Mean DICE Score | $0.65 \pm 0.06$ | $0.82 \pm 0.10$ |
| | Median DICE Score | $0.76 \pm 0.05$ | $0.84 \pm 0.04$ |
|  | T-Test vs Test Dataset; p-value |  | 0.068174 |

In the case of PHE, the median DSC was 0.71 (±0.02), reflecting a good agreement with the reference standard, p-value of 0.75. The volume difference for PHE was slightly higher at 12% (±9%), with an absolute volume difference of 0.57 (±1.84) ml. Corresponding Bland-Altman Analysis confirmed a trend where the difference increased as the mean volume exceeded approximately 100 ml, suggesting to overestimate larger volumes relative to the ground truth and is supported by illustrative cases provided in the **Supplementary Figure 1, D-E**. Nontheless, the Pearson correlation coefficient was strong at 0.92, paired with a strong correlation between automated and ground truth volumes in the scatter plot analysis in **Figure 4, A**. Detection accuracy was also high with 98% and a sensitivity of 99%.

The network achieved a median DSC of 0.68 (±0.09) for IVH, slightly below the scores for ICH and PHE, due in part to instances of unrecognized IVHs or false identifications, as indicated by the occurrence of ’0’ scores. **Supplementary Figure 2** demonstrated that misclassifications occurred predominantly in cases with anatomical overlap and morphological similarities to ICH or structures resembling IVH due to mass effect related changes in the anatomy. Despite lower DSCs, the volume difference was -1% (±10%), with an absolute volume difference of -0.87 (±0.83) ml, and a Pearson correlation of 0.99, indicating a high consistency in volume quantification. Upon assessing IVH with a volume threshold of > 1 ml, the mean DSC improved to 0.65 (±0.06) and the median DSC to 0.76 (±0.05) with the observed differences from the reference standard no longer reaching statistical significance, p-value of 0.07. This refinement was also reflected in the Bland-Altman plot where cases below 10 ml of volumes had the most outliers as shown in **Figure 3, A**. Overall, sensitivities and specifities remained high up to an IVH volume of 0.2 ml as detailed in the **Supplementary Table 2** and **Supplementary Figure 3**.

### Performance across different anatomical contexts

**Table 3** presents performance metrics by anatomical location, with **Figures 5-6** depicting the network’s effectiveness, particularly high precision for ICH and PHE. While IVH segmentation varied, with lower DSC in complex regions like the brainstem/pons, volume estimation correlations for IVH remained high across most regions.

**Table 3:** Localiaztion Specific Performance Evaluation: Segmentation and Volume Estimation Metrics Across different Locations in the Test Dataset. *Legend:* Performance metrics (1) for segmentation evaluation, including Dice score analysis, represented through mean and median values, along with pairwise comparison among various datasets; (2) volume estimation accuracy, utilizing both relative and absolute volume discrepancies, complemented by a Pearson correlation assessment between actual ground truth (GT) volumes and those predicted by the AI; and (3) the efficacy of detection across all data subjects and for those above 1 mL. Additional segmentation metrics for IVH exceeding 1 mL are specifically delineated in a separate subsection. Intracerebral hemorrhage (ICH), perihematomal edema (PHE) and intraventricular hemorrhage (IVH).

| Lesion Type | Performance Metrics | Lobar<br>N= 80 | Deep<br>N= 80 | Brainstem<br>N= 9 | Cerebellum<br>N= 20 |
| --- | --- | --- | --- | --- | --- |
| ICH Metrics | Mean DICE Score | $0.87 \pm 0.03$ | $0.88 \pm 0.03$ | $0.65 \pm 0.17$ | $0.82 \pm 0.06$ |
| | Median DICE Score | $0.90 \pm 0.01$ | $0.92 \pm 0.01$ | $0.70 \pm 0.29$ | $0.87 \pm 0.05$ |
|  | T-Test vs Lobar; p-value | - | 0.78762 | <0,0001 | 0.10145 |
| | Volume Difference in % | $-1\% \pm 8\%$ | $-2\% \pm 3\%$ | $104\% \pm 161\%$ | $17\% \pm 31\%$ |
| | Absolute Difference in ml | $-1.23 \pm 2.71$ | $-0.55 \pm 0.71$ | $1.75 \pm 3.57$ | $-0.26 \pm 1.13$ |
|  | Pearson Correlation Volume GT vs AI | 0.95 | 0.99 | 0.76 | 0.98 |
| | Detection Accuracy | $99\% \pm 2\%$ | $99\% \pm 2\%$ | $100\% \pm 0\%$ | $100\% \pm 0\%$ |
| | Detection Sensitivity | $99\% \pm 2\%$ | $100\% \pm 0\%$ | $100\% \pm 0\%$ | $100\% \pm 0\%$ |
| | Detection Specificity | NA | $0\% \pm 0\%$ | NA | NA |
| | Detection Accuracy Threshold of 1 ml | $97\% \pm 4\%$ | $96\% \pm 4\%$ | $78\% \pm 28\%$ | $100\% \pm 0\%$ |
| | Detection Sensitivity Threshold of 1 ml | $97\% \pm 3\%$ | $97\% \pm 4\%$ | $78\% \pm 28\%$ | $100\% \pm 0\%$ |
| | Detection Specificity Threshold of 1 ml | NA | $0\% \pm 0\%$ | NA | NA |
| PHE Metrics | Mean DICE Score | $0.69 \pm 0.03$ | $0.68 \pm 0.03$ | $0.49 \pm 0.12$ | $0.61 \pm 0.07$ |
| | Median DICE Score | $0.71 \pm 0.02$ | $0.72 \pm 0.02$ | $0.50 \pm 0.18$ | $0.65 \pm 0.08$ |
|  | T-Test vs Lobar; p-value | - | 0.86315 | 0.00014 | 0.03346 |
| | Volume Difference in % | $12\% \pm 10\%$ | $1\% \pm 8\%$ | $88\% \pm 143\%$ | $22\% \pm 22\%$ |
| | Absolute Difference in ml | $2.75 \pm 3.71$ | $-1.64 \pm 2.22$ | $1.82 \pm 2.96$ | $0.74 \pm 2.43$ |
|  | Pearson Correlation Volume GT vs AI | 0.9 | 0.93 | 0.8 | 0.94 |
| | Detection Accuracy | $99\% \pm 3\%$ | $98\% \pm 3\%$ | $100\% \pm 0\%$ | $95\% \pm 10\%$ |
| | Detection Sensitivity | $99\% \pm 3\%$ | $100\% \pm 0\%$ | $100\% \pm 0\%$ | $100\% \pm 0\%$ |
| | Detection Specificity | NA | $0\% \pm 0\%$ | NA | $0\% \pm 0\%$ |
| | Detection Accuracy Threshold of 1 ml | $99\% \pm 2\%$ | $97\% \pm 3\%$ | $78\% \pm 28\%$ | $95\% \pm 10\%$ |
| | Detection Sensitivity Threshold of 1 ml | $99\% \pm 2\%$ | $100\% \pm 0\%$ | $78\% \pm 28\%$ | $100\% \pm 0\%$ |
| | Detection Specificity Threshold of 1 ml | NA | $0\% \pm 0\%$ | NA | $0\% \pm 0\%$ |
| IVH Metrics | Mean DICE Score | $0.46 \pm 0.11$ | $0.61 \pm 0.10$ | $0.26 \pm 0.21$ | $0.39 \pm 0.19$ |
| | Median DICE Score | $0.62 \pm 0.2$ | $0.79 \pm 0.07$ | $0.22 \pm 0.29$ | $0.33 \pm 0.49$ |
|  | Dice Score T-Test vs Lobar p-value | - | 0.045641 | 0.23261 | 0.45215 |
| | Volume Difference in % | $-1\% \pm 17\%$ | $-3\% \pm 10\%$ | $-24\% \pm 82\%$ | $26\% \pm 46\%$ |
| | Absolute Difference in ml | $-2.53 \pm 4.73$ | $-0.91 \pm 1.26$ | $-5.53 \pm 5.89$ | $-0.30 \pm 1.82$ |
|  | Pearson Correlation Volume GT vs AI | 0.98 | 0.99 | 0.45 | 0.95 |
| | Detection Accuracy | $81\% \pm 9\%$ | $86\% \pm 8\%$ | $88\% \pm 22\%$ | $65\% \pm 22\%$ |
| | Detection Sensitivity | $96\% \pm 7\%$ | $98\% \pm 5\%$ | $100\% \pm 0\%$ | $100\% \pm 0\%$ |
| | Detection Specificity | $72\% \pm 13\%$ | $74\% \pm 15\%$ | $79\% \pm 36\%$ | $36\% \pm 30\%$ |
| | Detection Accuracy Threshold of 1 ml | $92\% \pm 6\%$ | $91\% \pm 6\%$ | $89\% \pm 22\%$ | $80\% \pm 19\%$ |
| | Detection Sensitivity Threshold of 1 ml | $90\% \pm 11\%$ | $88\% \pm 10\%$ | $75\% \pm 43\%$ | $78\% \pm 28\%$ |
| | Detection Specificity Threshold of 1 ml | $94\% \pm 7\%$ | $95\% \pm 7\%$ | $100\% \pm 0\%$ | $82\% \pm 24\%$ |
| IVH Threshold Metrics (> 1ml) | Mean DICE Score | $0.62 \pm 0.09$ | $0.72 \pm 0.08$ | $0.33 \pm 0.21$ | $0.56 \pm 0.19$ |
| | Median DICE Score | $0.71 \pm 0.07$ | $0.82 \pm 0.04$ | $0.32 \pm 0.29$ | $0.65 \pm 0.28$ |
|  | Dice Score T-Test vs Lobar p-value | - | 0.02072112 | 0.13046 | 0.6709 |

### Performance across concurrent hemorrhage types

The subgroup analysis, detailed in **Supplementary Table 3** and **Supplementary Figure 4**, revealed no significant impact of concurrent hemorrhages on ICH segmentation accuracy, with the network maintaining high DSC values. Illustrations in **Supplementary Figure 1**C-D and minimal correlation between hemorrhage type and DSC underscore the network’s consistency.

### Evaluation across different independent validation sets

**Table 4** compares the network’s performance across validation sets, with DSC box plot comparisons to ground truth in **Supplementary Figure 5**.

**Table 4:** Evaluation of nnU-Net Framework: Lesion Segmentation, Volume Estimation, and Detection Accuracy Across Validation Sets. *Legend:* Performance metrics (1) for segmentation evaluation, including Dice score analysis, represented through mean and median values, along with pairwise comparison among various datasets; (2) volume estimation accuracy, utilizing both relative and absolute volume discrepancies, complemented by a Pearson correlation assessment between actual ground truth (GT) volumes and those predicted by the AI; and (3) the efficacy of detection across all data subjects and for those above 1 mL. Additional segmentation metrics for IVH exceeding 1 mL are specifically delineated in a separate subsection. Intracerebral hemorrhage (ICH), perihematomal edema (PHE) and intraventricular hemorrhage (IVH).

| Lesion Type | Performance Metrics | Spontaneous ICH Case Set |  | Diverse ICH Case Set |
| --- | --- | --- | --- | --- |
|  |  | Validation Set #1 | Validation Set #2 | Validation Set #3 |
| ICH Metrics | Mean DICE Score | 0.82 +- 0.03 | 0.81 +- 0.02 | 0.81 +- 0.04 |
|  | Median DICE Score | 0.88 +- 0.01 | 0.85 +- 0.01 | 0.90 +- 0.01 |
|  | T-Test vs Test Dataset; p-value | 0.0056844 | 0.00025 | 0.001626 |
|  | Volume Difference in % | -18% +- 3% | 21% +- 6% | 48% +- 74% |
|  | Absolute Difference in ml | -6.11 +- 1.25 | 6.84 +- 1.41 | 0.89 +- 0.66 |
|  | Pearson Correlation Volume GT vs AI | 0.99 | 0.97 | 0.98 |
|  | Detection Accuracy | 97% +- 2% | 99% +- 2% | 96% +- 3% |
|  | Detection Sensitivity | 97% +- 3% | 99% +- 2% | 96% +- 3% |
|  | Detection Specificity | NA | NA | NA |
|  | Detection Accuracy Threshold of 1 ml | 89% +- 4% | 97% +- 3% | 81% +- 6% |
|  | Detection Sensitivity Threshold of 1 ml | 89% +- 5% | 97% +- 3% | 81% +- 6% |
|  | Detection Specificity Threshold of 1 ml | NA | NA | NA |
| PHE Metrics | Mean DICE Score | 0.61 +- 0.02 | 0.57 +- 0.02 | 0.57 +- 0.03 |
|  | Median DICE Score | 0.65 +- 0.01 | 0.61 +- 0.02 | 0.66 +- 0.03 |
|  | T-Test vs Test Dataset; p-value | <0.001 | <0.001 | <0.001 |
|  | Volume Difference in % | 55% +- 21% | -11% +- 5% | -26% +- 10% |
|  | Absolute Difference in ml | 10.78 +- 2.86 | -6.65 +- 2.04 | -4.01 +- 1.10 |
|  | Pearson Correlation Volume GT vs AI | 0.86 | 0.93 | 0.93 |
| | Detection Accuracy | 97% ± 3% | 98% ± 2% | 95% ± 3% |
| | Detection Sensitivity | 97% ± 3% | 98% ± 2% | 96% ± 3% |
|  | Detection Specificity | NA | NA | NA |
| | Detection Accuracy Threshold of 1 ml | 93% ± 4% | 95% ± 3% | 85% ± 5% |
| | Detection Sensitivity Threshold of 1 ml | 93% ± 4% | 96% ± 3% | 86% ± 5% |
|  | Detection Specificity Threshold of 1 ml | NA | NA | NA |
| IVH Metrics | Mean DICE Score | 0.61 ± 0.06 | 0.46 ± 0.06 | 0.53 ± 0.10 |
| | Median DICE Score | 0.78 ± 0.04 | 0.62 ± 0.18 | 0.70 ± 0.16 |
|  | T-Test vs Test Dataset; p-value | 0.0210991 | 0.34598 | 0.73521 |
| | Volume Difference in % | -1% ± 10% | -10% ± 10% | -10% ± 10% |
| | Absolute Difference in ml | -0.87 ± 0.83 | 0.19 ± 1.44 | 0.19 ± 1.44 |
|  | Pearson Correlation Volume GT vs AI | 0.99 | 0.91 | 0.99 |
| | Detection Accuracy | 89% ± 4% | 78% ± 6% | 92% ± 4% |
| | Detection Sensitivity | 94% ± 4% | 79% ± 8% | 97% ± 5% |
| | Detection Specificity | 83% ± 8% | 76% ± 11% | 90% ± 5% |
| | Detection Accuracy Threshold of 1 ml | 89% ± 4% | 72% ± 7% | 94% ± 4% |
| | Detection Sensitivity Threshold of 1 ml | 81% ± 7% | 63% ± 9% | 76% ± 14% |
| | Detection Specificity Threshold of 1 ml | 99% ± 2% | 90% ± 8% | 99% ± 2% |
| IVH Threshold Metrics (> 1ml) | Mean DICE Score | 0.49 ± 0.07 | 0.68 ± 0.05 | 0.69 ± 0.08 |
| | Median DICE Score | 0.67 ± 0.13 | 0.80 ± 0.03 | 0.80 ± 0.08 |
|  | T-Test vs Test Dataset; p-value | 0.72394 | 0.001785 | 0.26366 |

Segmentation precision for ICH across the validation sets was notably consistent, with median DSC of 0.88 (±0.01) for Set #1, 0.85 (±0.01) for Set #2, and 0.90 (±0.01) for Set #3, also when compared to the test set’s median DSC of 0.91 (±0.01). For PHE, the validation sets showed a median DSC of 0.65 (±0.01) for Set #1, 0.61 (±0.02) for Set #2, and 0.66 (±0.03) for Set #3, which were all closely aligned with the test set’s median of 0.67 (±0.03). For IVH, the median DSCs were consistent in Sets #2 and #3, both reporting 0.80 (±0.03 for Set #2 and ±0.08 for Set #3), slightly surpassing the score in Set #1 and the test set (median of 0.67 (±0.13 and median of 0.76 (±0.05), respectively). In agreement with these findings,

The network showed high Pearson correlations for ICH volume across validation sets (0.97-0.99), with strong volume estimation in PHE and IVH, despite some variability. In Set #1, PHE volume differed by 55% (±21%), with a 0.86 Pearson correlation, while Sets #2 and #3 maintained correlations above 0.90, comparable to the test set, as shown in **Figures 3 and 4**. IVH estimations revealed variability for smaller volumes but improved correlation with larger volumes, and overall, Bland-Altman plots confirmed the network’s robust volume estimations across lesion types and data sets.

### Analysis time

The trained model demonstrated an average processing time of 18.2 seconds per case for automatic segmentation and volume analysis, in contrast to an mean time of 18.01 minutes (± SD 20.47 minutes) for manual segmentation.

## Discussion

Our study presents a comprehensive analysis of an advanced biomedical segmentation network for the segmentation of ICH, PHE, and IVH. We utilized a substantial training cohort of 775 patients from three different European centers and validated its performance across three different validation sets that encompassed varied clinical and imaging contexts.

The network exhibited excellent segmentation precision for ICH, achieving high DSC even in challenging imaging scenarios marked by irregular shapes, multifocal bleeding sites, or instances where various types of hemorrhage co-occurred. Despite these strengths, a consistent pattern of volume underestimation for cases exceeding 100 mL was noted, mirroring findings reported by Sharrok et al.^28^ This underscores a potential area for model enhancement, particularly for scenarios involving hyperacute hemorrhage. In such cases, the hypodensity from non-clotted blood might resemble the appearance of perihematomal edema (PHE), as depicted in **Supplementary Figure 1**, highlighting the need for advanced differentiation capabilities in the model’s design.

While PHE segmentation yielded the lowest DSC compared to the two other lesion types, its alignment with the inter-rater ground truth—the benchmark for our study—validated the trustworthiness of our findings. Moreover, the high Pearson correlation coefficient underscored the network’s effectiveness in volumetric quantification over exact contour replication, which, from a clinical perspective, holds more significance than the precise replication of lesion contours.^2,29^

In the segmentation of IVH, our network’s precision declined with smaller IVH volumes and in differentiating between similar conditions or complex anatomical structures, as indicated by reduced DSC scores and detailed in **Supplementary Figure 3**. It is well-documented that the DSC exhibits a pronounced sensitivity to volume, where inaccuracies are magnified in smaller hemorrhagic volumes.^30^ A minor misclassification of voxels can lead to a significantly lower DSC in small-scale IVH. This impact is exemplified by the fact that a 1 mL error in a 40 mL hemorrhage results in a 97.5 DSC, whereas the same error in a 4 mL hemorrhage yields a DSC of 0.75. Addressing the DSC’s volume sensitivity, our network applied a threshold refinement for lesions over 1 mL—a clinically recognized benchmark— resulting in enhanced segmentation precision.^23^ This benchmark aligns with standards set by leading FDA-approved software for intracranial hyperdensities.^24^ Importantly, our analysis demonstrated that the network’s sensitivity and specificity remained above 90% for IVH volumes as small as 0.2 mL, sizes traditionally considered clinically non-significant. These findings pave the way for future studies to further refine measurement uncertainty and explore the clinical implications of detecting smaller volume hemorrhages. The variabilities in IVH segmentation within complex anatomical structures, such as the brainstem, pons, and cerebellum, are likely due to the lower incidence of hemorrhages in these regions, mirroring their epidemiological rarity. Despite these complexities, often compounded by strong imaging artifacts and patient motion, our network still maintained commendable performance, and producing reliable results as illustrated in the **Supplementary Figure 6**.

The comparative analysis across different independent validation sets revealed a commendable consistency in the segmentation performance, with minimal variance from the test set’s results. Notably, the network’s performance on Validation Set #3, which included a broader range of ICH etiologies, showcased its robust generalizability even to previously unseen conditions. This study is, to the best of our knowledge, the first to illustrate such capabilities, setting it apart from the studies reported by Zhao et al. and Kok et al. for the automated segmentation of ICH, IVH, and PHE.^6,7^ Kok et al.’s study utilized a large dataset from the international multicenter TICH-2 trial (Tranexamic Acid for Hyperacute Primary Intracerebral Haemorrhage) to assess various segmentation models.^7^ Kok et al. achieved a median DSC of 0.92 for ICH, closely matched by our network’s median DSC of 0.91, demonstrating parallel precision. In addressing the complexities of IVH segmentation outlined above, Kok et al.’s research also encountered similar challenges. Their nnU-Net model was specifically adapted to tackle the issue of class imbalance regarding IVH within the dataset. They managed these difficulties by implementing a focal loss function, which significantly improved the segmentation of IVH. This adaptation echoes the broader struggle and innovative approaches in the field to enhance IVH segmentation accuracy. Zhao et al.’s single-center study featured a modest cohort of 380 patients.^6^ Despite the smaller sample size, their findings showcased excellent agreement in ICH and IVH segmentation, with DSC of 0.92 and 0.79, respectively, along with an impressive case processing time of 15 seconds, findings that align closely with our own. Furthermore, our study expands on this by including follow-up data in Validation Set #2 to reinforce the temporal accuracy of our model’s predictions, providing an improvement over the approaches of both Kok et al. and Zhao et al., who analyzed solely baseline CT scans.

While our study acknowledges minor segmentation errors in ground truth annotations, as exemplified by subtle IVH instances in **Supplementary Figure 2**, the robust inter-rater reliability documented in our previous work suggests these inaccuracies represent a negligible fraction of cases.^23,31^ Moreover, the retrospective design of our study may introduce biases that necessitate prospective validation. Nonetheless, our multicenter cohort and the inclusion of three independent validation sets support the robustness of our findings. From a clinical perspective, the implementation of this model in clinical practice may be constrained by the computational resources required for processing as well as the ’black box’ nature of the employed deep learning model which may limit interpretability, a critical factor in clinical decision-making.

Our study, leveraging a large European cohort, affirms its effectiveness in estimating ICH, PHE, and IVH volumes, an aspect more clinically crucial than perfectly delineating lesion contours. Validated against three independent sets, including diverse etiologies unseen by the network, the study underscores the model’s generalizability and robustness, further strengthened by incorporating data from scanners of four different manufacturers. The automatic method proved to be 98.32% faster than the manual approach. Our findings signal an encouraging shift towards the adoption of automated segmentation networks in clinical practice, offering significant gains in both efficiency and consistency. Nonetheless, the nuanced performance variances noted across varying volumes and anatomical contexts for IVH, together with the comparatively lower DSC scores for PHE, point to specific areas for targeted enhancement, setting the stage for advanced, precision-oriented clinical tools.

## Data Availability

The datasets that support the findings of our study are available upon reasonable request from the corresponding author. However prior approval of proposals may apply by our institution's data security management and a signed data sharing agreement will then be approved.

## Supplementary Material

### Supplement: 2.2 Data Collection and Patient Demographics

The study included patients who (1) were diagnosed with primary, spontaneous ICH; (2) were aged 18 or above; and (3) had baseline Non-Contrast Computed Tomography (NCCT) scans performed within the first 36 hours of onset or last known well (LSW) status. We excluded cases of (1) hemorrhages caused by trauma, vascular malformations, tumors, hemorrhagic conversion of ischemic strokes, or other non-primary ICH sources; (2) bleeding that was non-parenchymal (3) insufficient or inadequate imaging data. Table 1 summarizes the additional clinical information gathered from patient records, complemented by detailed imaging data derived from the analysis outlined below.

### Supplement: 2.4 Imaging Analysis and Image Quality Assessment

Images were anonymized and stored in Digital Imaging and Communications in Medicine (DICOM) format. Two-dimensional DICOM images were converted to a three-dimensional structure using the Neuroimaging Informatics Technology Initiative format (NIfTI), re-orienting the images if necessary and resampling to a slice thickness of 5 mm after performing quality assessment via instance number and slice distance checking, as well as filtering scans with few slices.^32^ Additional subjective quality assessment was performed for the presence of artifacts, slice missing, and low axial resolution. CT scans were analyzed for the presence of IVH and hemorrhage location, followed by the imaging annotation for the three different labels ICH, PHE, and IVH as described previously.^31^ Volume regions of interest (VOI) were manually derived for each label using region segmentation on each slice with a polygon tool using the ITK Snap software (version 3.8.0).^33^ Investigators were trained blinded to subject identity, clinical information as well as the other readers’ interpretations. This approach was validated by our previous study, which demonstrated outstanding interobserver reliability, as reflected by Intraclass Correlation Coefficients: 0.998 for ICH, 0.979 for IVH, and 0.886 for PHE. Notably, intrarater agreement was even higher, with coefficients between 0.98 to 0.99, underscoring the consistency and reproducibility of our segmentation process.^31^ An expert in ICH imaging (J.N.) reviewed all images for IVH presence, evaluated the segmentation accuracy of the three labels, and made corrections where necessary. In the final step, we implemented a refined subjective assessment method by adopting a batched strategy, utilizing an HTML table layout, to display all scans in a web browser which were reviewed by an experienced investigator (J.N). Any scans that did not meet the necessary criteria were excluded, ultimately leading to the exclusion of a total of N=111 images after conducting both objective and subjective quality assessments as described.^32^

### Supplement: 2.5 Deep Learning Network Architecture

Many different architectural designs exist to implement semantic segmentation. In this study, we employed the nnU-Net framework, a state-of-the art automated segmentation tool specifically engineered for biomedical image segmentation.^13^ This method distinguishes itself by dynamically adjusting to each new dataset, thereby avoiding the need for static hyperparameters.^14^ Therefore, interrelated hyperparameters are grouped into three distinct groups: first, the blueprint parameters, which are held constant across all datasets; second, the inferred parameters, which are tailored dynamically to meet the unique demands of each dataset; and third, the empirical parameters, which cannot be predefined and must instead be derived from the data itself.^14^ Besides that, the nnU-Net network architecture mirrors that of the U-Net, employing an encoder-decoder structure enhanced with skip connections and an output stride of 1, as elaborated in **Figure 1**. The encoder captures the essential contextual information necessary to distinguish among the three classes: ICH, PHE, and IVH. The decoder then progressively restores this information to the original image resolution, merging the upsampled contextual data from the preceding layers with the finer-resolution feature maps via skip connections.^14^ The network’s depth, determined by the input size, encompassed seven stages of downsampling and upsampling.^13^ Architecturally, the network adopts a standard configuration with two blocks at each resolution level for both encoder and decoder. Each block comprises a sequence of convolution, instance normalization, and leaky ReLU nonlinearity, establishing a balance between complexity and performance.^34^

#### Supplement: 2.6.1 Training and Testing

Subjects from Charité Berlin, University Medical Center-Hamburg-Eppendorf, Germany, and IRCCS Mondino Foundation in Pavia, Italy were combined for training and optimization of hyperparameters. We allocated 20% of the data for testing purposes. The remaining 80% of the dataset was used in a 5-fold cross-validation framework. In each fold, we divided this 80% into five equal parts. Four of these parts, constituting 64% of the total dataset, were used for training, while the fifth part was reserved for validation. This approach ensures that each segment of the dataset is used for both training and validation, enhancing the robustness of our model. For the final prediction, all five folds, each representing a separately trained instance of our nnU-Net model, were utilized collectively. It is important to note that the 20% of the dataset designated for testing was kept entirely separate and was not used in either the training or validation phases. To analyze performance during training and validation, we calculated the average test results from each fold for ICH, PHE, and IVH. Subsequently, we compared the predicted masks and the time required for segmentation of these labels, juxtaposed with their respective ground truth masks and the time expended for manual segmentation in the test images for each fold.

#### Supplement: 2.6.2 Validation

To reinforce the robustness of our segmentation network, we further substantiated our results by evaluating the network’s performance across three different validation datasets.

1. <u>Internal Validation Set:</u> Data from the validation set #1 consisted from University Medical Center Hamburg-Eppendorf that facilitated the reproducibility of our results.
2. <u>External Validation Set:</u> Data from the validation set #2 came from an independent institution, University Hospital Munster, Germany, which had not contributed prior datasets to the initial training. This dataset enabled a more extensive validation of our network’s generalizability and accuracy, independent of the specific local imaging protocols and CT scanner manufacturers.
3. <u>Diverse Validation Set:</u> Data from the validation set #3 of Charité Berlin were not limited to cases of spontaneous ICH but included a more generalized representation encompassing various ICH subtypes encompassing hypertensive bleedings, and those cases associated with cerebral amyloid angiopathy, oral anticoagulation, and vascular malformations as detailed in our previous study Nawabi et al^12^. Such a comprehensive cohort selection, allowed for a robust assessment of our network across a broad spectrum of ICH presentations, thereby enhancing the generalizability of our findings.

### Supplement: 2.7 Training and Loss functions

Models were trained for 1000 epochs with a learning rate of 0.01, and a batch size of 12 images. Dice loss complemented with the categorical cross-entropy loss were used as loss function.^15^ The categorial Cross-Entropy Loss is a Softmax activation plus a Cross-Entropy loss and considered as a popular loss function for multi-classification problems, which takes the output probabilities (P) and measures the distance from the truth values according to the following formula:

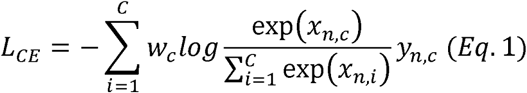

where x is the input, y is the target, w is the weight, and C is the number of classes.

The Dice Similarity Coefficient (DSC) quantifies the volumetric overlap between the segmented outcome and the ground truth.^20^ It calculates this by comparing set A, which consists of the foreground voxels in the ground truth, with set B, the equivalent foreground voxels in the segmentation output, which is defined as follows:

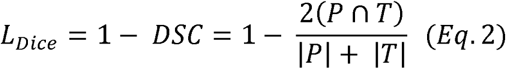

where ∩ is the intersection and *(P* ∩ *T)* represents the spatial overlap between *P* and *T*. *T* denotes the ground truth segmentation and *P* denotes the predicted segmentation. | *P* | and | *T* | represent the areas of *P* and T, respectively.

### Supplement: 2.8 Performance Evaluation Metrics

#### 2.8.1 Segmentation

The performance of each network for the segmentation estimations of ICH, PHE, and IVH was evaluated on test data after the training and validation process as well as in the validation sets. To quantify the quality of the segemtation estimations, we calculated the DSC as an established validation metric of spatial overlap index.^20^ The DSC measured the spatial overlap between two segmentations, the predicted region (P) and the verified ground truth (T) region as elaborated in Equation 2. Paired samples t-test was used for comparative analyses of DSC values derived from repeated ground truth segmentations and the validation sets against those obtained from the network’s predictions.

#### Supplement: 2.8.2 Volume Estimation

To assess the precision of volume estimations, the Absolute Volume Error (AVE) was computed to reflect the deviation of predicted volumes from the ground truth as well as absolute relative volume difference (ARVD), as explicated in Equation 3 and 4.^21,22^ The ARVD is the absolute volume difference calculated relative to the volume of the ground truth. Furthermore, we examined the relationship between predicted and ground truth volumes using Pearson’s correlation coefficient (R).^21^ The results were reported as point estimates with the corresponding standard deviation, delineated by the double standard deviation from 1000 bootstrap resamples.

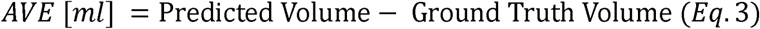

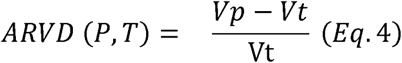

Where V_p_ represents the predicted volume and V_t_ the true (ground truth) volume. A perfect score of 0 indicates that the objects have identical volume, but does not necessarily mean that the prostates are well aligned spatially.^22^

#### Supplement: 2.8.3 Detection

To ascertain the performance of our automated detection system, we systematically evaluated its accuracy, sensitivity, and specificity. The latter metrics were calculated by benchmarking the automated detection of ICH, PHE, and IVH against the control group with healthy individuals which comprised N=50 NCCT scans. The mathematical formulations corresponding to the specified metrics are detailed in Equations 5 through 7.

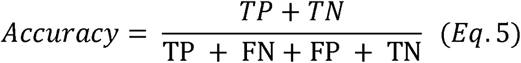

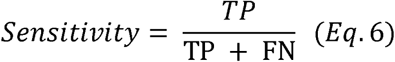

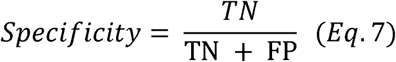

Where TP: True Positive, TN: True Negative, FP: False Positive, FN: False Negative, TP represents the number of samples with a presence of IVH correctly classified as IVH positive, while FN represents the number of IVH samples wrongly classified to negative IVH class. Similarly, TN represents the number of samples without IVH presence correctly classified to as IVH negative cases while FP represents the number of samples without IVH presence wrongly classified as IVH positive. The described method for classification accuracy applies uniformly across other lesion types such as ICH and PHE, where TP, TN, FP, and FN correspondingly indicate correctly or incorrectly classified cases with or without the presence of the respective lesion.

#### Supplement: 2.8.4 Subgroup Analysis for ICH and IVH

The network’s capability in identifying lesions for ICH, PHE, and IVH was assessed separately focusing on cases with volumes exceeding 1 ml, as this volume threshold addresses a methodological gap prevalent in both the current literature and FDA-approved hemorrhage detection software.^23,24^ This assessment was also extended to IVH segmentation evaluations as this particular volume threshold has been recognized as a marker of clinical relevance. ^37^

A second subgroup analysis was performed to evaluate the network’s accuracy in segmenting ICH when concurrent secondary intra- and extracranial bleedings, such as SAH, SDH, or EDH, were present. This involved comparing DSC for ICH lesions both with and without accompanying hemorrhages, and assessing the correlation between bleeding type and segmentation accuracy using Pearson’s correlation coefficient.

### Supplement: 2.9 A Web-based User-Interface for Radiological Report

The final step involved the development of a web-based user-interface (UI), the ICH-Viewer, for visualization of segemtatations and reporting of volumes. The front-end UI was written using using the Cornerstone.js - JavaScript library.^26^ Cornerstone is used by default for image retrieval, decoding, and rendering of DICOM images.^27^ It can leverage CPU- and GPU-based rendering to display medical imaging data sets. These services are orchestrated through a back-end realized using Python scripts. In our implementation, our UI is created with Flask, a Python based front end development tool that updates hypertext modeling language (HTML) files.^36^ Information is captured through an HTML form supplemented with JavaScript, which is the processed through a decision support module to provide predictions for the different volumes and presence of IVH. The results are then presented in a web-friendly format which can be copied and pasted into a radiologist’s reporting software. The application includes a robust image viewer alongside the display of segmentation maps created with the network alongside the generated report including volume quantification of ICH, PHE, IVH as well as information on the presence of IVH. **Figure 2** shows the UI with a representative snapshot of the ICH-Viewer Interface

---

**Supplementary Table 1:** Specifications of Computed Tomography (CT) Scanners Across Participating Sites. **Legend:** Listed specifications for Computed Tomography (CT) scanners used at different medical institutions. Specifications include the kilovoltage peak (KVP) in kilovolts (kV), the tube current in milliamperes (mA), the slice reconstruction thickness in millimeters (mm), and the in-plane resolution also in millimeters (mm).

| CT Scanner |  | KVP [KV] | I [mA] | slice reconstruction [mm] | in-plane resolution [mm] |
| --- | --- | --- | --- | --- | --- |
| **Charité Berlin, Germany** |  |  |  |  |  |
| Toshiba CT Aquillion Prime | Canon Medical Systems Corporat | 120 | 270 | < 5.0 | < 0.5 |
| Toshiba Aquilion ONE ViSION | Canon Medical Systems Corporat | 120 | 250 | < 5.0 | < 0.5 |
| Toshiba Aquilion PRIME 160 | Canon Medical Systems Corporat | 120 | 280 | < 5.0 | < 0.5 |
| Toshiba CT Aquillion Prime ONE | Canon Medical Systems Corporat | 120 | 250 | < 5.0 | < 0.5 |
| LightSpeed VCT Ultra 64 | GE Healthcare - Global Headqua | 120 | Dose-Modulation (100-450) | < 5.0 | 0.5 |
| Revolution EVO | GE Healthcare | 120 | Dose-Modulation (100-365) | < 5.0 | 0.625 |
| Revolution GSI | GE Healthcare | 120 | Dose-Modulation (100-365) | < 5.0 | 0.625 |
| Revolution CT | GE Healthcare | 120 | Dose-Modulation (200-400) | < 5.0 | 0.625 |
| Aquilion PRIME 160 with Fluoro | Canon Medical | 120 | 270 | < 5.0 | 0.5 |
|  | Systems<br>Corporat |  |  |  |  |
| Aquilion<br>PRIME 160 | Canon<br>Medical<br>Systems<br>GmbH | 120 | 180 | < 5.0 | 0.5 |
| Aquilion ONE<br>Prism | Canon<br>Medical<br>Systems<br>GmbH | 120 | 350 | < 5.0 | 0.5 |
| **University Medical Center Hamburg-Eppendorf, Germany** |  |  |  |  |  |
| Philips iCT<br>256 | Philips<br>Healthcare | 120 | 280-320 | < 5.0 | 0.5 |
| **University Hospital Munster, Germany** |  |  |  |  |  |
| Siemens<br>Somatom<br>Definition<br>Flash | Siemens<br>Deutschland<br>Healthcare | 120 | 280 | < 5.0 | 0.5 |
| **IRCCS Mondino Foundation, Italy** |  |  |  |  |  |
| Toshiba<br>Aquilion<br>Prime | Canon<br>Medical<br>Systems<br>GmbH | 120 | 250 | < 5.0 | 0.5 |

**Supplementary Table 2:** Sensitivity and Specifity for IVH Detection across different Datasets. **Legend:** Sensitivity and specificity rates for IVH detection plotted for volumes above 0.2 mL across various datasets

|  | Treshhold of 0.2 ml |  |
| --- | --- | --- |
| Data Set | Sensitivity | Specificity |
| Test Set | 0.94 (0.88-0.99) | 0.91 (0.85-0.96) |
| Validation Set #1 | 0.88 (0.82-0.94) | 0.96 (0.92-1) |
| Validation Set #1 | 0.73 (0.64-0.81) | 0.91 (0.83-0.99) |
| Validation Set #3 | 1 (1-1) | 0.96 (0.93-1) |

**Supplementary Table 3:** Comparative Analysis of Segmentation Precision in Parenchymal Hemorrhages with and without other coexisting Intracranial Hemorrhages Types. *Legend:* Mean and Median Dice Similarity Coefficients for Segmentation of Intracerebral Hemorrhage with or without Extension into Subarachnoid, Epidural, or Subdural Spaces, Including Associated Correlation Coefficients. DICE, Dice Similarity Coefficients; EDH, epidural hematoma; SAH, subarachnoid hemorrhage; SDH, subdural hematoma.

| **SAH** | **With** | **Without** | **p-value** |
| --- | --- | --- | --- |
| Mean DICE Score | 0.90 ± 0.04 | 0.86 ± 0.14 | 0.3067 |
| Median DICE Score | 0.90 ± 0.05 | 0.91 ± 0.07 |  |
| Pearson Correlation Coefficient |  | 0.075 |  |

| **SDH and/or EDH** | **With** | **Without** | **p-value** |
| --- | --- | --- | --- |
| Mean DICE Score | 0.87 ± 0.05 | 0.87 ± 0.14 | 0.71858 |
| Median DICE Score | 0.91 ± 0.06 | 0.90 ± 0.07 |  |
| Pearson Correlation Coefficient |  | 0.027 |  |

**Supplementary Figure 1:**
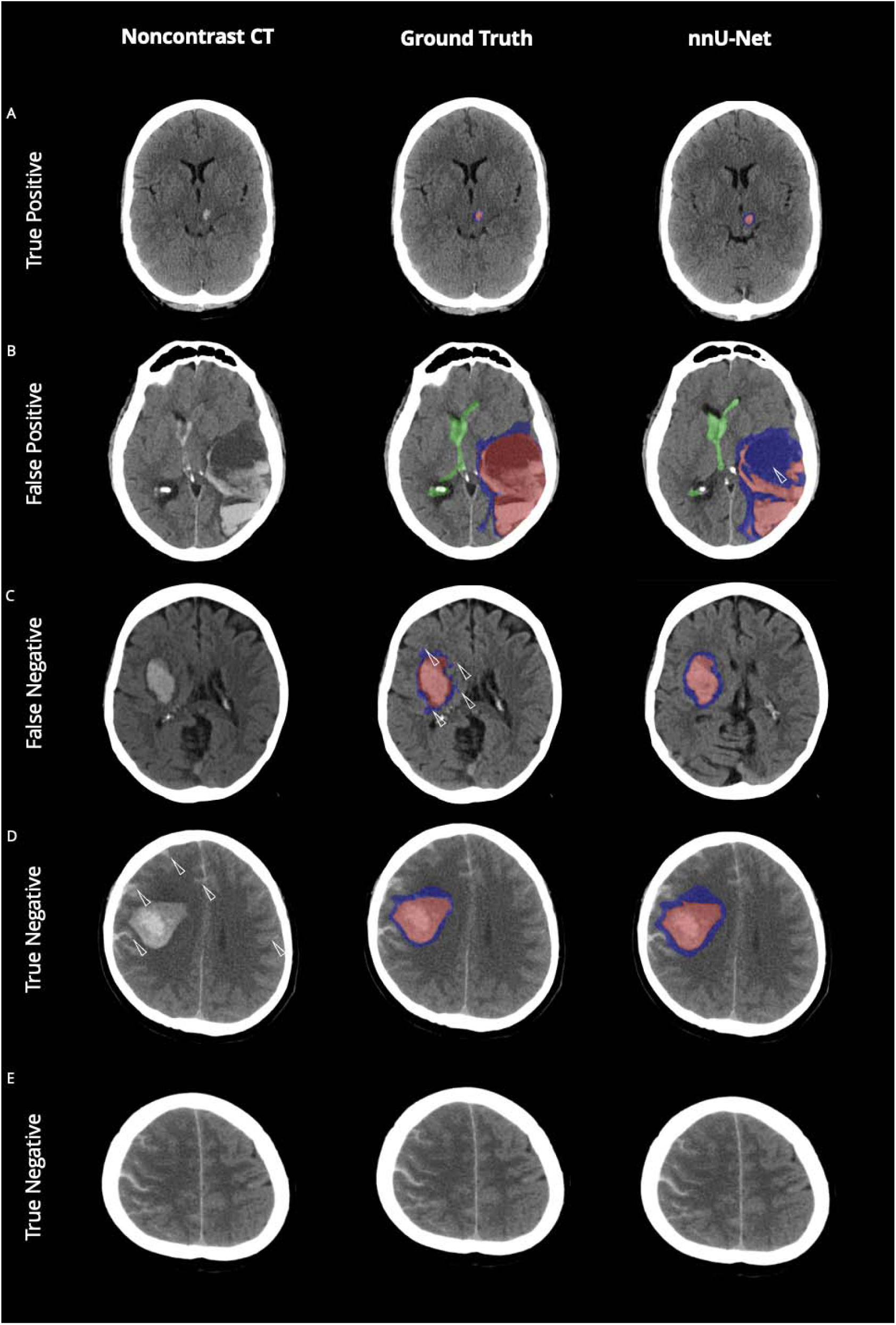
Evaluation of Intracerebral and Perihematomal Edema Predictions - A Visual Comparison of Model Accuracy and Misclassifications. *Legend:* Noncontrast Computed Tomography scans with the original data (left column), ground truth segmentations (mid column) and the network’s predicted segmentations (nnU-Net, right column). A: High accuracy in identifying small intracerebral hemorrhage (ICH; < 5 ml). B: Misclassification error in identifying a hypodense portion of the hemorrhage as perihematomal edema (PHE) potentially attributed to the similar densities as the latter as indicated by the white arrow. C: Correct classification of PHE; however showing that the network tented to smooth contours relatively to the ground truth. D-E: High accuracy in identidying subarachnoid hemorrhage extension (SAHE; white arrows) along the cortical fissures as non-parenchymal ICH. The color-coding is as follows: intracerebral hemorrhage (ICH) is highlighted in red; and perihematomal edema (PHE) is depicted in blue.

**Supplementary Figure 2:**
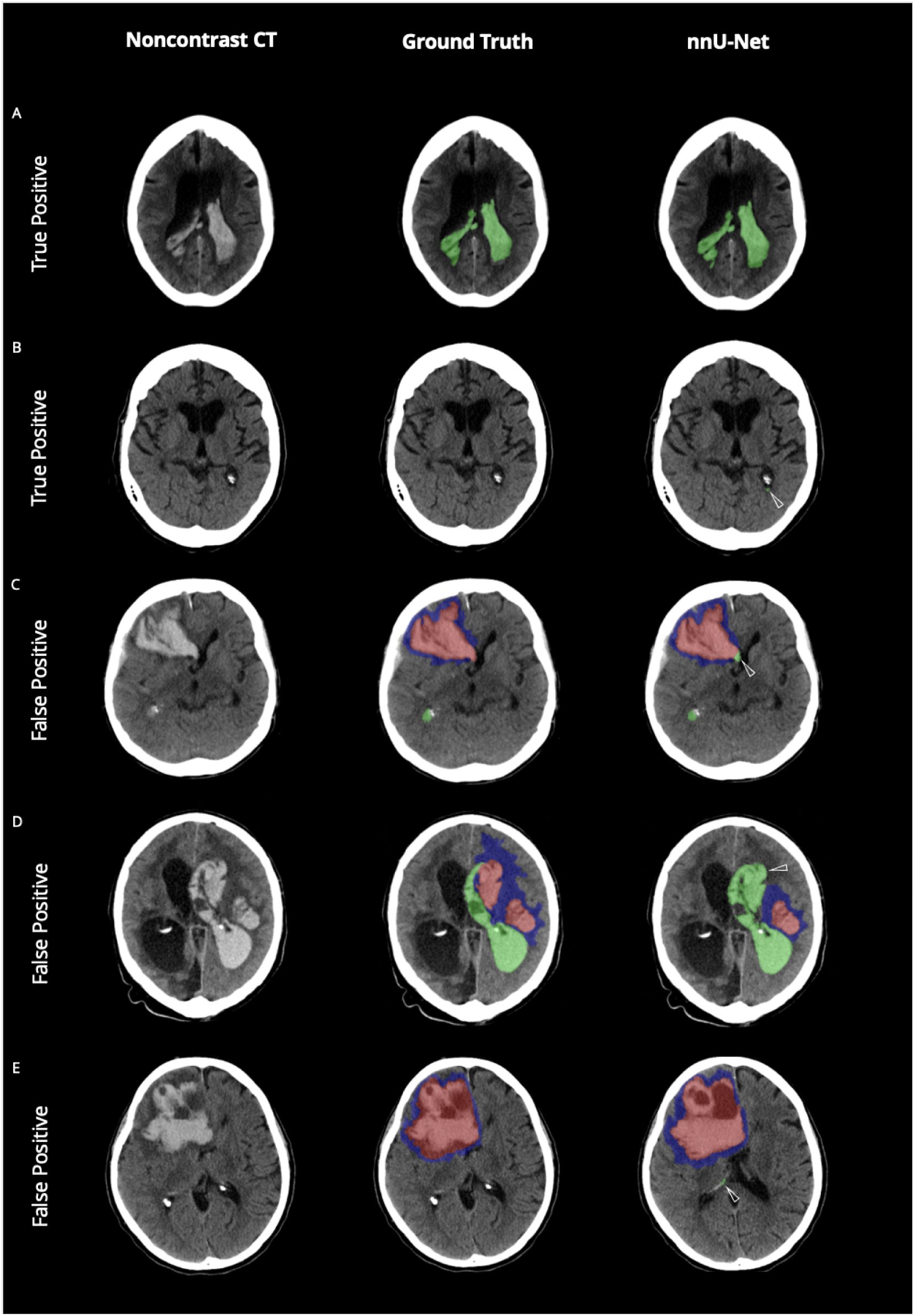
Evaluation of Intraventricular Hemorrhage Predictions - A Visual Comparison of Model Accuracy and Misclassifications. *Legend:* Noncontrast Computed Tomography scans with the original data (left column), ground truth segmentations (mid column) and the network’s predicted segmentations (nnU-Net, right column). A: High accuracy in the prediction of a large and heterogenous intraventricular hemorrhage (IVH) extension into the lateral ventricles. B: Correct classification of a small IVH in the left lateral ventricle juxtaposed to the missed classification in the ground truth. C-D: Missclassification of ICH as IVH as indicated by the white arrow in case of overlapping anatomical context and IVH-like shape. E: Missclassification of IVH as indicated by white arrow in case of IVH-shape like formation of the choroid plexus due to ICH induced mass effect. The color-coding is as follows: intracerebral hemorrhage (ICH) is highlighted in red; intraventricular hemorrhage extension (IVH) is marked in green; and perihematomal edema (PHE) is depicted in blue.

**Supplementary Figure 3:**
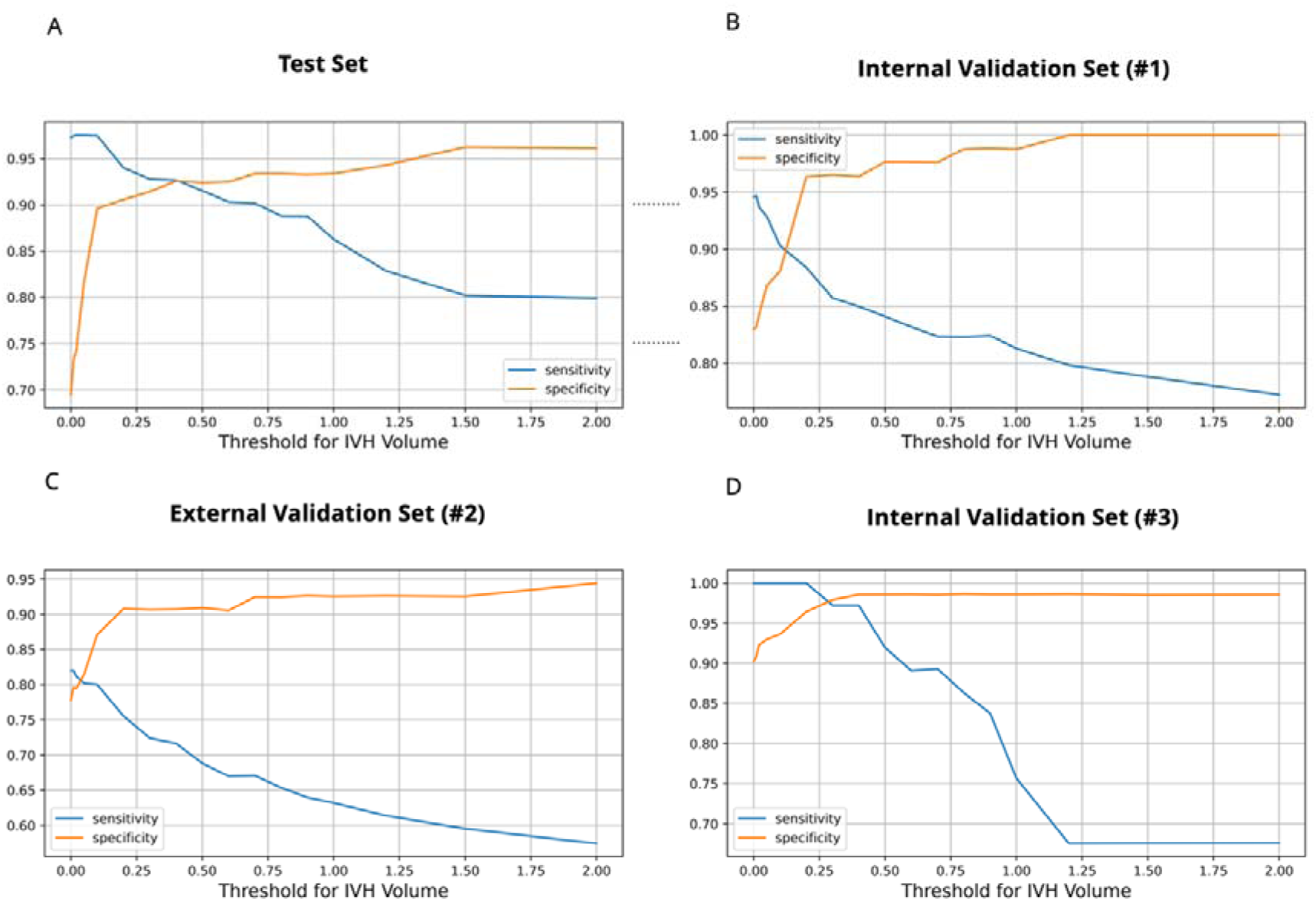
Volume-dependant Variances in Sensitivity and Specificity for IVH Detection. **Legend:** This figure illustrates the relationship between IVH volume and the sensitivity (blue) and specificity (orange) of IVH detection across four datasets: the test set (A) and the three validation sets (B-D). The graphs display how sensitivity and specificity rates vary with increasing IVH volume thresholds, indicating the model’s optimal metrics at between 0.2-0.25 ml.

**Supplementary Figure 4:**
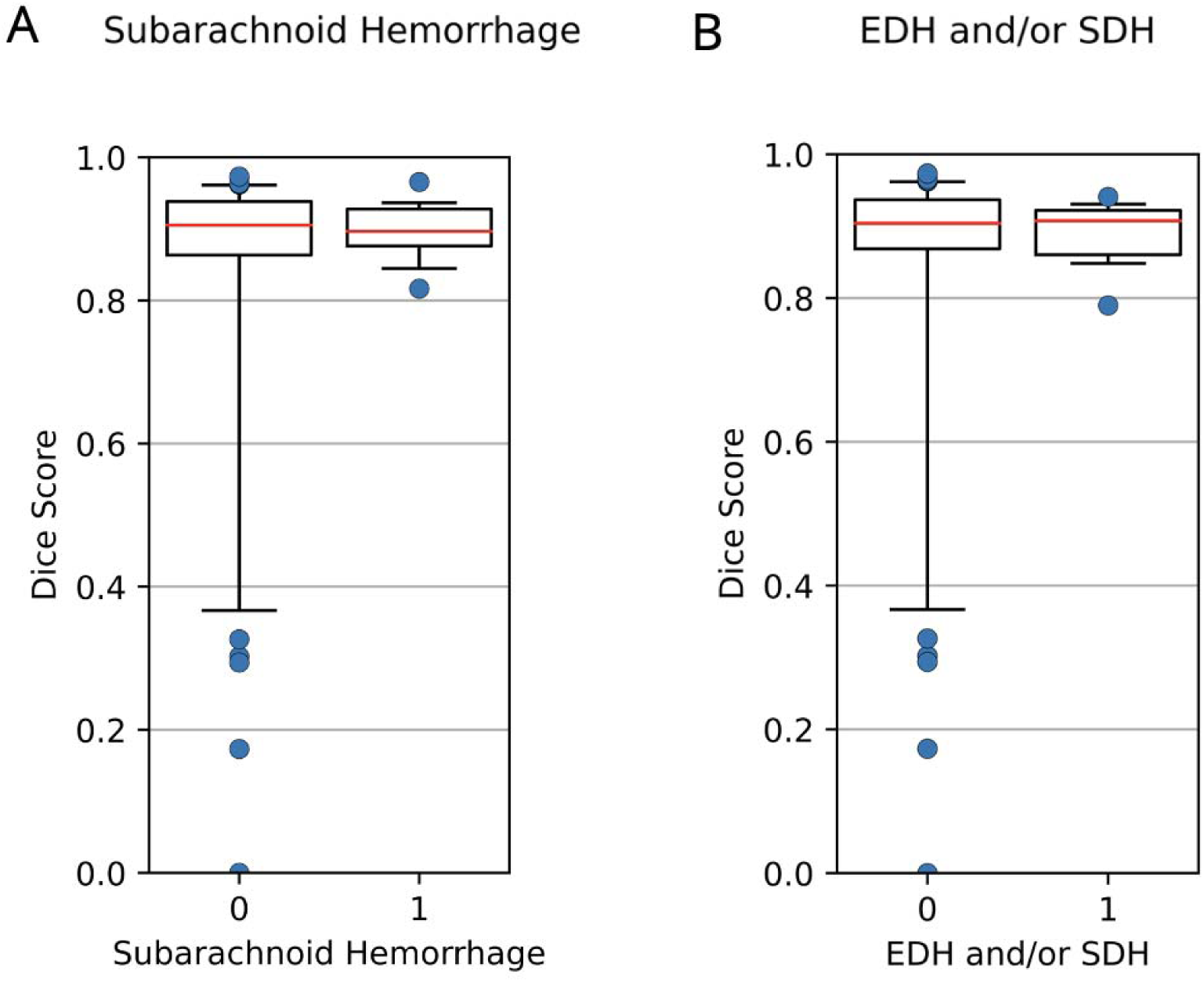
Segmentation Performance for ICH with with Associated Secondary Bleedings. *Legend:* Series of boxplots illustrating the comparative accuracy of the network’s segmentation predictions for cases of intracerebral hemorrhage (ICH) with concurrent secondary bleedings. A: Dice similaatiry coefficient (DSC) for segmentations when subarachnoid hemorrhage is present. B: DSC for cases with either epidural (EDH) or subdural hematoma (SDH). Each boxplot represents the distribution of Dice scores, with the central line indicating the median, the box denoting the interquartile range (IQR), and the whiskers extending to the most extreme data points that are no more than 1.5 times the IQR from the upper and lower quartiles. Outliers are not individually plotted to preserve the clarity of the visualization.

**Supplementary Figure 5:**
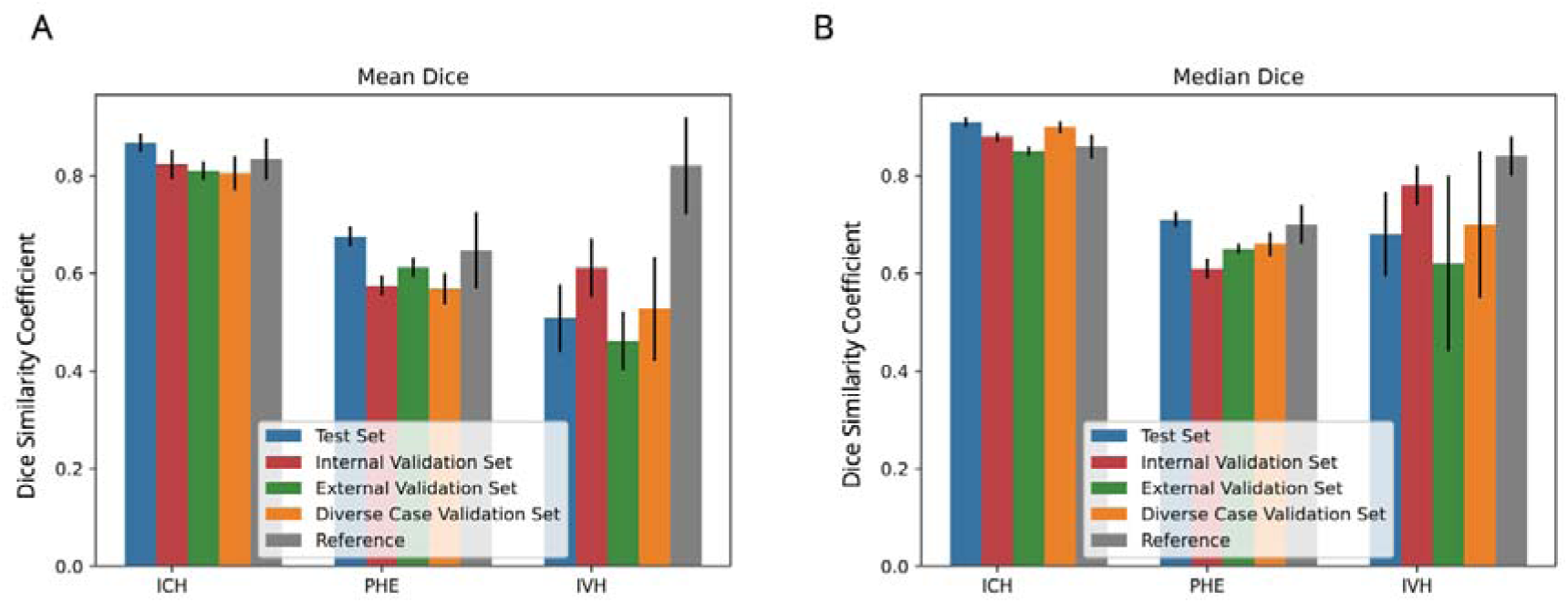
Comparative Analysis of Segmentation Accuracy Across Test and different Validation Sets for ICH, PHE, and IVH Labels. **Legend:** Boxplot comparison of DICE Scores for segmentation accuracy across different sets. Panel A presents mean DICE Scores, while Panel B shows median values. The metrics are compared for intracerebral hemorrhage (ICH), perihematomal edema (PHE), and intraventricular hemorrhage (IVH). Color coding represents the test set (blue), homogeneous validation sets with only spontaneous ICH cases (internal validation in red, external validation in green), and a heterogeneous validation set with a wide spectrum of ICH causes (orange). DICE scores from manual segmentations are indicated in grey for reference. Whiskers extend to the most extreme data points within 1.5 times the 95% CI from the quartiles, and outliers are omitted from individual plotting to maintain visual clarity.

**Supplementary Figure 6:**
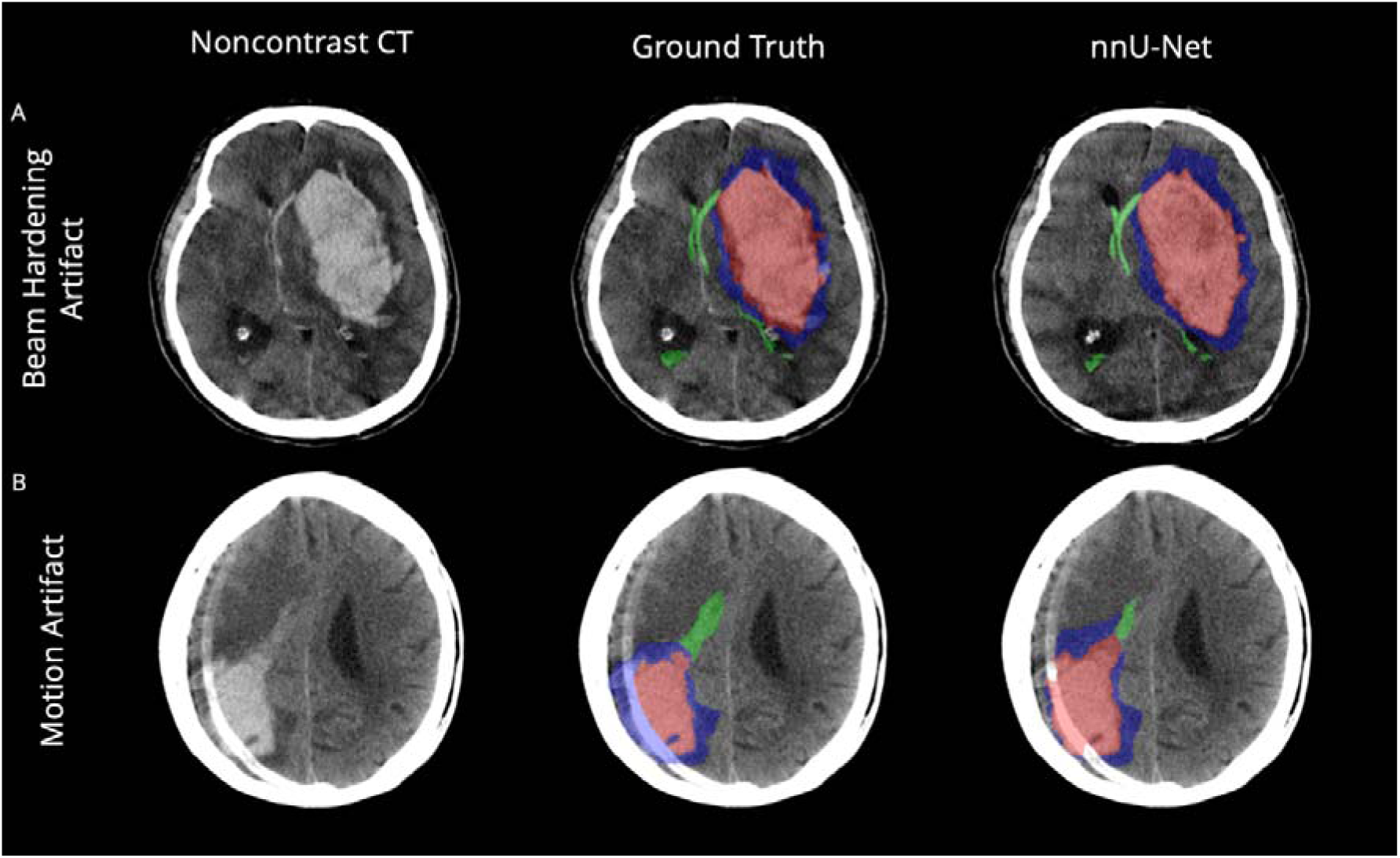
Evaluation of Imaging Artifaacts - A Visual Comparison of Model Accuracy. *Legend:* Noncontrast Computed Tomography scans with the original data (left column), ground truth segmentations (mid column) and the network’s predicted segmentations (nnU-Net, right column). A: High accuracy in identifying all lesions in case with beam hardening artifact B: Moderate accuracy in identifying all lesions in case with motion artifact. The color-coding is as follows: intracerebral hemorrhage (ICH) is highlighted in red; and perihematomal edema (PHE) is depicted in blue.

